# Racial/Ethnic Differences in Neuropsychological Test Performance in Frontotemporal Degeneration

**DOI:** 10.1101/2025.01.06.25320069

**Authors:** Melanie A. Matyi, Emma Rhodes, Sheina Emrani, Hannah A. Jin, David J. Irwin, Corey T. McMillan, Lauren Massimo

**Affiliations:** 3700 Hamilton Walk, Richards Medical Laboratories, Penn Frontotemporal Degeneration Center, Department of Neurology, School of Medicine, University of Pennsylvania, Philadelphia, PA 19104, USA; 677 Huntington Avenue, Harvard T.H. Chan School of Public Health, Department of Biostatistics, Boston, MA 02115, USA; 418 Curie Boulevard, Fagin Hall, Department of Biobehavioral Health Sciences, School of Nursing, University of Pennsylvania, Philadelphia, PA 19104, USA

**Author notes:** Corresponding Author: Lauren Massimo, PhD, 3700 Hamilton Walk, B603 Richards Medical Laboratories, Philadelphia, PA 19104, USA.

**Keywords:** frontotemporal degeneration, cognition, minoritized groups

## Abstract

**BACKGROUND:** Racial and/or ethnic differences in neuropsychological test performance are understudied in frontotemporal degeneration (FTD) but their identification is critical to identifying ways to improve care of representative FTD populations.

**METHODS:** Differences in cognitive scores between Black and Hispanic relative to White participants were assessed with the sequential addition of potential contributing factors. Group differences in the likelihood of impairment status in cognitive test performance were also evaluated.

**RESULTS:** Minoritized individuals had lower scores and/or greater likelihood of impairment on measures of lexical retrieval, processing speed, cognitive flexibility, and working memory but not global cognition, verbal recall, attention, and category fluency. Addition of factors attenuated group differences.

**DISCUSSION:** Racial/ethnic differences on neuropsychological tests used in diagnosis and monitoring of FTD were substantially attenuated when accounting for potential contributing factors. To address these differences in FTD, future efforts must increase representative research participation of patients and understand social determinants of health.

## Background

Frontotemporal degeneration (FTD) is a common cause of young-onset dementia that results in progressive deterioration in executive functioning, language and social comportment. Limited research has examined the epidemiology of FTD across minoritized groups but population-based studies from across the globe indicate that individuals of all races and ethnicities are affected by FTD^1^. Although limited information on the prevalence of FTD across racial/ethnic groups exists, all-cause dementia including Alzheimer’s Disease (AD) are more common in minoritized groups^2^. The greater prevalence of dementia in minoritized groups is due, may be due in part, to these individuals performing worse on neuropsychological tests that are sensitive to cultural bias, thus reducing diagnostic accuracy^3,4^. We previously demonstrated differences in the clinical presentation of Asian and Black groups compared to White individuals diagnosed with FTD (behavioral variant FTD [bvFTD] or primary progressive aphasia [PPA])^5^. In particular, Black individuals, compared to White individuals, exhibited greater functional impairment and distinct neuropsychiatric symptom profiles at initial National Alzheimer’s Coordinating Center (NACC) visits. However, cognitive performance differences between racial/ethnic groups have not been examined in individuals with FTD.

Extant research has compared neuropsychological test performance of Black and Hispanic individuals to White individuals, but this has been largely reported in non-clinical/community samples of older adults^6,7^, all-cause dementia^8,9^ or AD^10–12^. On average, minoritized individuals score lower on traditional neuropsychological tests than White individuals in both dementia and cognitively normal samples and across the lifespan^6,13^. Education quality ^14^ and access to resources that promote better health outcomes, including socio-economic status, nutrition, and insurance are thought to drive observed differences in performance on neuropsychological tests^15–17^, in addition to evidence that traditional neuropsychological test are sensitive to cultural bias^18^.

Previous research demonstrates poorer neuropsychological test performance among minoritized individuals diagnosed with dementia, particularly AD, relative to White individuals^8,9,12^, and differences in the clinical presentation of FTD in Black compared to White individuals^5^. However, performance differences on neuropsychological testing between racial/ethnic groups with an FTD diagnosis have not been reported. Thus, we sought to identify potential contributors to differences in cognitive performance across racial/ethnic groups of individuals with an FTD diagnosis from a large national cohort, the National Alzheimer’s Coordinating Center (NACC). To achieve these aims, we conducted linear regressions first adding racial/ethnic group to estimate the total difference and then sequentially adding potential contributing factors. This method allows for assessment of the extent to which potential contributing factors attenuate observed racial/ethnic differences in performance. Second, we conducted binary logistic regression to assess if total racial/ethnic group differences translated into a greater likelihood of Black and Hispanic individuals being classified as impaired, relative to White individuals, controlling for disease severity.

## Methods

Data were obtained from the National Alzheimer’s Coordinating Center (NACC), a public dataset established by the National Institute on Aging that collects standardized clinical and neuropathological data^19^. Data used in this study is from the December 2023 data freeze which included data collected from 2005 to 2023 and from 39 ADRCs. Participants were included in this study if 1) they had an etiologic diagnosis of primary progressive aphasia (PPA; NACC variable NACCPPA) or behavioral variant FTD (bvFTD; NACC variable NACCBVFT), 2) self-identified as Black, Hispanic or White, 3) had complete demographic information (i.e., age, sex, education, and onset of cognitive decline), 4) had a Clinical Dementia Rating (CDR®) Dementia Staging Instrument Global score greater than 0, and 5) completed neuropsychological testing in their primary language as detailed in Figure S1. The first NACC visit in which data was collected for one of the ten neuropsychological tests was examined to capture the earliest cognitive performance. For most participants, the initial NACC visit was examined (mean = 1.33; SD = 1.00) with the initial visit being the most often examined visit for Black (76.6%), Hispanic (91.1%) and White (82.3%) individuals.

Only (57/103) (55%) of minoritized participants completed all ten neuropsychological tests examined in the current study at a single visit. In order to maximize the sample size for minoritized groups, individuals were included in analyses if they completed at least one of the neuropsychological tests. To reduce bias, the same criterion was used for determining the sample for the White group. Additionally, we used propensity matching to ensure that the composition of the White group was similar to that of the Black and Hispanic groups. Consistent with a previous study^5^, each member of the minoritized groups was matched to 1 or more White individuals. Matching was performed by calculating the distance between participants via generalized linear model with a probit link. A caliper of 0.2 on the standardized distance between participants was used for matching such that any 2 participants with a standardized distance greater than 0.2 on covariates are not matched^20^. Covariates used for matching included age at visit, CDR global, education, sex and the pattern of missing data. We matched on the pattern of missing data because individuals missing data for at least one neuropsychological test tended to perform worse on other neuropsychological tests. To account for potential systematic differences in specifically which tests were missing, groups were matched on the pattern of missing data. For example, if a Black participant had all tests except digit span forward (DSF), then they were matched to a White participant who completed all tests except DSF and had no more than 0.2 standardized distance on other covariates (age, CDR, education, sex). Because participants were included even if they did not complete all tests, different subsets of Black (n=28-54), Hispanic (n=40-75) and White (n=1,212-1,715) participants were included in analyses for specific neuropsychological tests. See Table 1 for cognitive measures and their corresponding sample sizes.

**Table 1.** Sample Characteristics.

| Characteristic | Overall<br>N = 1,819<sup>1</sup> | White<br>N = 1,716<sup>1</sup> | Black<br>N = 47<sup>1</sup> | Hispanic<br>N = 56<sup>1</sup> | p-<br>value<sup>2</sup> |
| --- | --- | --- | --- | --- | --- |
| **Age (years)** | 65.18 (9.31) | 65.16 (9.30) | 65.87 (9.43) | 65.04 (9.43) | 0.8 |
| **Sex** |  |  |  |  | 0.015 |
| Male | 1,085 (60%) | 1,036 (60%) | 19 (40%) | 30 (54%) |  |
| Female | 734 (40%) | 680 (40%) | 28 (60%) | 26 (46%) |  |
| **Education (years)** | 15.62 (2.74) | 15.69 (2.71) | 14.70 (2.89) | 14.34 (3.24) | <0.001 |
| **Disease duration (years)** | 4.70 (2.93) | 4.70 (2.93) | 4.60 (2.86) | 4.57 (3.04) | >0.9 |
| **Global CDR** |  |  |  |  | 0.026 |
| Very mild | 851 (47%) | 814 (47%) | 20 (43%) | 17 (30%) |  |
| Mild | 674 (37%) | 636 (37%) | 15 (32%) | 23 (41%) |  |
| Moderate | 220 (12%) | 200 (12%) | 8 (17%) | 12 (21%) |  |
| Severe | 74 (4.1%) | 66 (3.8%) | 4 (8.5%) | 4 (7.1%) |  |
| **NACC visit number** | 1.33 (1.00) | 1.34 (1.02) | 1.30 (0.62) | 1.23 (0.85) | 0.2 |
| **Stroke** | 21 (1.2%) | 19 (1.1%) | 2 (4.3%) | 0 (0%) | 0.2 |
| **Diabetes** | 166 (9.1%) | 149 (8.7%) | 7 (15%) | 10 (18%) | 0.027 |
| **Hypertension** | 700 (38%) | 641 (37%) | 27 (57%) | 32 (57%) | <0.001 |
| **Clinical phenotype** |  |  |  |  | 0.6 |
| bvFTD | 981 (54%) | 922 (54%) | 25 (53%) | 34 (61%) |  |
| PPA | 838 (46%) | 794 (46%) | 22 (47%) | 22 (39%) |  |
| **UDS version** |  |  |  |  | 0.4 |
| 1 | 407 (22%) | 383 (22%) | 14 (30%) | 10 (18%) |  |
| 2 | 858 (47%) | 816 (48%) | 18 (38%) | 24 (43%) |  |
| 3 | 554 (30%) | 517 (30%) | 15 (32%) | 22 (39%) |  |
| Global cognition: MMSE/MoCA | 1,817 (99.9%) | 1,715 (99.9%) | 47 (100%) | 55 (98%) | 0.1 |
| Immediate recall: Logical<br>Memory/Craft Story 21 | 1,676 (92%) | 1,590 (93%) | 37 (79%) | 49 (88%) | <0.001 |
| Delayed recall: Logical Memory/Craft<br>Story 21 | 1,671 (92%) | 1,587 (92%) | 37 (79%) | 47 (84%) | <0.001 |
| Attention: Digit/Number Span Forward | 1,699 (93%) | 1,609 (94%) | 39 (83%) | 51 (91%) | 0.015 |
| Working memory: Digit/Number Span<br>Backward | 1,695 (93%) | 1,606 (94%) | 39 (83%) | 50 (89%) | 0.017 |
| Category fluency: Animals | 1,709 (94%) | 1,618 (94%) | 39 (83%) | 52 (93%) | 0.009 |
| Category fluency: Vegetables | 1,681 (92%) | 1,594 (93%) | 38 (81%) | 49 (88%) | 0.008 |
| Processing speed: TMT A | 1,589 (87%) | 1,509 (88%) | 36 (77%) | 44 (79%) | 0.013 |
| Cognitive flexibility: TMT B | 1,271 (70%) | 1,212 (71%) | 27 (57%) | 32 (57%) | 0.017 |
| Lexical retrieval: Naming<br>(BNT/MINT) | 1,670 (92%) | 1,585 (92%) | 37 (79%) | 48 (86%) | 0.001 |

<sup>1</sup>Mean (SD); n (%)<sup>2</sup>Kruskal-Wallis rank sum test; Pearson's Chi-squared test; Fisher's exact test
Note. bvFTD=behavioral variant frontotemporal dementia, PPA=primary progressive aphasia, UDS = uniform data set, MMSE=Mini Mental State Exam, MoCA=Montreal Cognitive Assessment, TMT=Trail Making Test, BNT=Boston Naming Test, MINT=Multilingual Naming Test.

Analyses were performed with R Studio version 4.3.2 (R Foundation). Symptom duration was calculated as age at the onset of cognitive decline until age at the visit date. To determine the age at the onset of cognitive decline, the NACC variable DECAGE, which is the clinician assessed age at which cognitive decline began, was used. Participants with symptom duration 4SDs above the mean (231.84 months) were excluded. Continuous demographic variables are summarized with mean and SD and categorical demographic variables are summarized with frequencies. Kruskal-Wallis rank sum tests, Pearson’s Chi-squared tests, and Fisher’s exact test (with Monte Carlo simulation using 2000 replicates to estimate p-values) were used as appropriate to assess group differences.

Data were collected across NACC Uniform Data Set (UDS) versions 1-3. We examined four neuropsychological tests that were collected across UDS versions 1-3 (i.e., category-guided fluency Animals and Vegetables and Trail Making Tests A [TMT-A] and B [TMT-B]). Between NACC UDS Versions 2 and 3.0, four proprietary tests from the neuropsychological battery were replaced with non-proprietary tests (see Table 2). Consistent with a prior cross-walk study, the original and replacement tests demonstrate good correlation^21^, and thus, data can be examined across UDS versions. Thus, a total of ten neuropsychological assessments were examined as listed in Table 1. In the current study, this cross-walk study was used to compute Version 3 scores that equate to Versions 1 and 2 for the purpose of examining differences in cognitive performance by racial/ethnic group. We also applied age, sex, and education adjusted norms derived for UDS Versions 1 and 2^19,22^, and for UDS Version 3^23^ to the corresponding raw data for the purpose of examining differences in impairment rates on cognitive tests by racial/ethnic group.

**Table 2.**
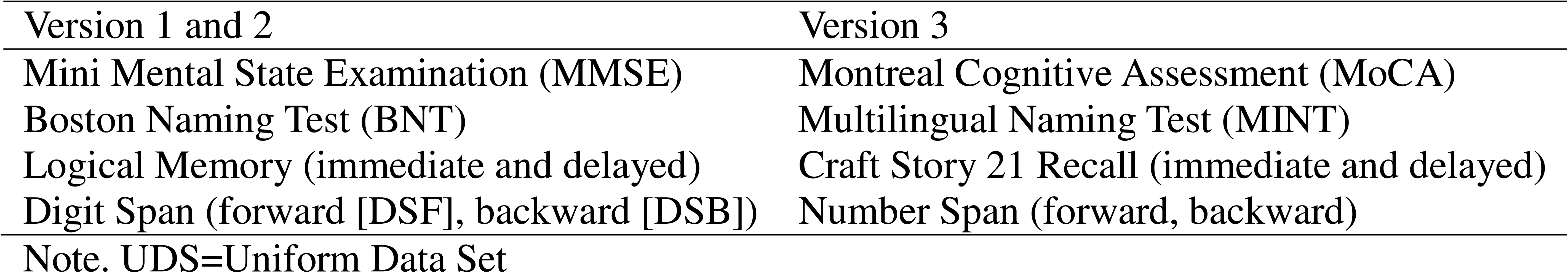
Neuropsychological Tests by UDS Versions.

| Version 1 and 2 | Version 3 |
| --- | --- |
| Mini Mental State Examination (MMSE) | Montreal Cognitive Assessment (MoCA) |
| Boston Naming Test (BNT) | Multilingual Naming Test (MINT) |
| Logical Memory (immediate and delayed) | Craft Story 21 Recall (immediate and delayed) |
| Digit Span (forward [DSF], backward [DSB]) | Number Span (forward, backward) |

Note. UDS=Uniform Data Set

Linear regression examined the association between race/ethnicity and cognitive scores (after applying cross-walk transformations to UDS Version 3 data). We implemented a series of linear regressions, first assessing the relationship of neuropsychological test performance with racial/ethnic group alone to examine the total racial/ethnic difference in performance. We then sequentially added CDR Global as a measure of disease severity (reference = 0.5 [very mild]), age, sex (reference = male), years of education, vascular comorbidities including recent stroke, presence of diabetes and presence of hypertension, and lastly, a missing data indicator of whether any of the 10 cognitive tests were missing (reference = complete data). We report detailed regression results for all additive models in the Supplement (Tables S1-S10). For all categorical variables, the reference group was the largest group for that variable, and thus, the White group was used as the reference group. Stroke was considered present if the (co-) participant reported a recent/active stroke or the clinician assessed there to be a silent or symptomatic stroke. Diabetes was considered present if the (co-) participant reported presence of diabetes or the clinician assessed there to be diabetes at the NACC visit. Hypertension was considered present if the (co-) participant reported recent/active hypertension or the clinician assessed hypertension to be currently present at the NACC visit.

Next, we implemented binary logistic regression to examine group differences in the proportion of participants classified as impaired on a given test covarying for CDR Global, using the threshold of <= −1.5 standard deviations below the normative sample mean, consistent with prior studies^19,23^. For all analyses the CDR, rather than the FTLD-CDR, was used as a measure of disease severity as the FTLD-CDR was not available for UDS Versions 1 and 2 (70% of participants).

Missingness of neuropsychological testing data significantly contributed to regression models; therefore for each measure, we report missingness and reasons that were provided for missing data by racial/ethnic group. In the NACC UDS, missing data is coded as one of the following: cognitive/behavior problem, other problem, physical problem, verbal refusal, or not available. Group differences in missingness are reported in Table 1 and reasons for missing data are reported in Supplemental Figure S2.

### Supplementary Analyses

Within group analyses: To further understand potential contributors to the observed racial/ethnic differences in neuropsychological test performance, we conducted analyses within racial/ethnic group, testing the effect of age, education, sex, and vascular comorbidities on neuropsychological test performance accounting for disease severity. Results are reported in Supplemental Tables S11-12.

Effect of language: We conducted supplemental analyses of neuropsychological test performance among all individuals identifying as Black, Hispanic and White, regardless of primary language, and including Asian participants (see Supplemental Table S13). These analyses are identical to the regressions described above with the additional predictor of participant’s primary language. Results are reported in Supplemental Fig. S3 and Tables S14-23.

## Results

### Participant Characteristics

The final sample included 47 (2.58%) Black, 56 (3.08%) Hispanic and 1,716 (94.34%) White individuals. As seen in Table 1, participants from minoritized groups had a higher frequency of incomplete data with only 25 (53.19%) Black and 32 (57.14%) Hispanic participants having completed all ten tests examined, whereas 1,206 (70.23%) of White participants had complete data. Participant demographic characteristics by group are also reported in Table 1. After matching individuals from minoritized groups to one or more White individuals, age at visit, visit number, UDS form version, disease duration, clinical phenotype, and recent/current stroke did not differ between Black or Hispanic and White individuals. Even after matching, minoritized individuals had fewer years of education (Black: β = −0.98; SE = 0.40; p = .015; Hispanic: β = −1.35; SE = 0.37; p < .001) and Hispanic individuals had a greater proportion of moderate and severe CDR Global scores than White individuals (p = .018). Minoritized individuals were also more likely to report presence of hypertension (Black: β = 0.82; SE = 0.30; p = .006; Hispanic: β = 0.80; SE = 0.27; p = .003). Black individuals were more likely to be female (Black: β = 0.81; SE = 0.30; p = .007) than White individuals. Hispanic individuals were more likely to report a history of diabetes (β = 0.83; SE = 0.36; p = .021) than White individuals.

### Differences in cognitive test performance between minoritized and White groups

Regressing neuropsychological measures on race/ethnicity gives the total difference between White and Black and Hispanic participants in scaled units. We identified differences in global cognition, processing speed and lexical retrieval for both Black and Hispanic participants. The difference in Global Cognition (measured by the MMSE or MoCA) is −0.43 (*p* < .05) for Black and −0.33 (*p* < .05) for Hispanic participants. The difference in processing speed (measured by TMT-A) is 0.39 (*p* < .05) for Black and 0.47 (*p* < .05) for Hispanic participants. The difference in lexical retrieval (measured by the BNT or MINT) is −0.51 (*p* < .05) for Black and −0.43 (*p* < .05) for Hispanic participants. We identified a difference only for Black participants for working memory (measured by DSB; −0.40; *p* < .05) and cognitive flexibility (measured by TMT-B; 0.60; *p* < .05). We identified a difference only for Hispanic participants for immediate recall (−0.38; *p* < .05) and delayed recall (−0.31; *p* < .05). We identified no difference for attention (measured by DSF) and category fluency (measured by Animals and Vegetables) for Black and Hispanic participants. Note that for all tests except the trail making tests, a higher score is indicative of better performance. For each neuropsychological test, the total racial/ethnic difference (top row of each panel in Fig. 1 and first column of Tables S1-10), and the subsequent difference after adjusting for additional predictors (additional rows of each Fig. 1 panel and columns of Tables S1-S10).

**Figure 1.**
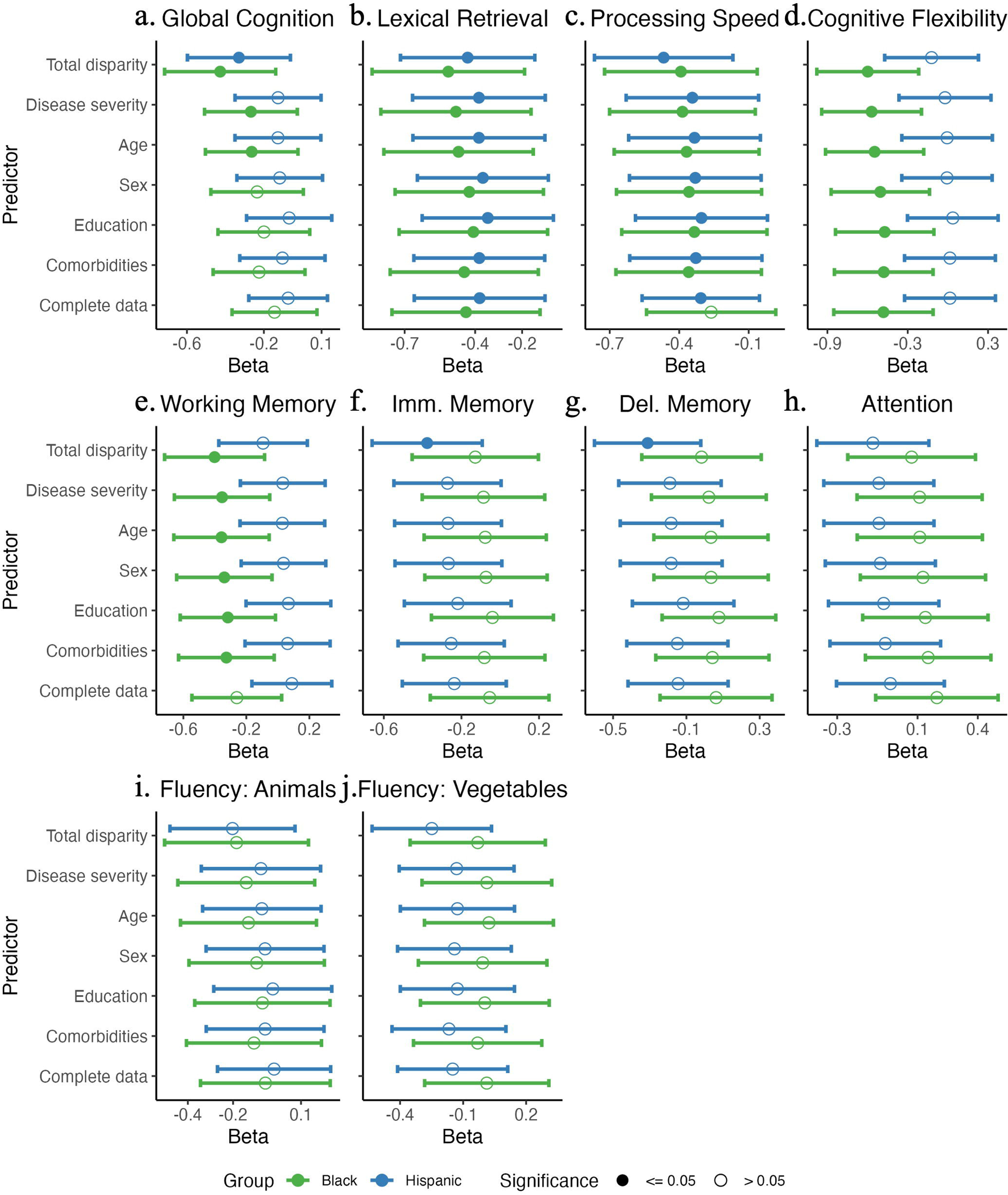
Estimated racial/ethnic disparities by linear regression models for each neuropsychological test examined. Coefficient terms for the difference adjusting for predictors listed on the y-axis and 95% confidence intervals are displayed. Coefficients and confidence intervals were multiplied by −1 for processing speed and cognitive flexibility so that the direction of effect (higher score reflects better cognition) is the same across all tests. Abbreviations: Imm. = immediate, Del. = delayed.

### Global Cognition (Fig 1a; Table S1)

The difference in global cognition is −0.43 (*p* < .05) for Black and −0.33 (*p* < .05) for Hispanic participants. As expected, greater disease severity, added next, was associated with lower global cognition. Additionally, disease severity attenuated the racial/ethnic difference to - 0.27 (*p* < .05) for Black and to −0.13 (*p* > .05) for Hispanic participants. Adding individual-level covariates shows that sex, education and, to a lesser extent, age, are significantly associated with global cognition and further attenuate the racial/ethnic difference in Global Cognition to non-significance for both Black (−0.20; *p* > .05) and Hispanic (−0.07; *p* > .05) participants. The presence of vascular comorbidities (stroke, diabetes and/or hypertension), added to the model next, did not further attenuate the racial/ethnic difference in global cognition. However, the absence of diabetes and hypertension is related to worse Global Cognition. The subsequent addition of data missingness (i.e., whether any of the 10 tests were missing) further reduced racial/ethnic differences in Global Cognition to −0.14 for Black and −0.07 for Hispanic participants. Participants missing at least one test had lower global cognition than those with complete data.

### Lexical Retrieval (Fig 1b; Table S2)

The total racial/ethnic difference in lexical retrieval, as measured by the BNT or MINT, is −0.51 (*p* < .05) for Black and −0.43 (*p* < .05) for Hispanic participants. As expected, greater disease severity was associated with worse lexical retrieval and attenuation of the total difference for Black (−0.48; *p* < .05) and Hispanic (−0.38; *p* < .05) participants. For individual-level covariates, age and sex, but not education, were related to lexical retrieval. All individual-level covariates (age, sex and education) attenuated the racial/ethnic difference for Black (−.41; *p* < .05) and Hispanic (−0.35; *p* < .05) participants. The presence of vascular comorbidities exacerbated the racial/ethnic difference for Black (−0.45; *p* < .05) and Hispanic (−0.38; *p* < .05) participants. Both absence of diabetes and hypertension were related to worse lexical retrieval. The missing data indicator was not related to lexical retrieval scores and did not attenuate the racial/ethnic differences in lexical retrieval. Thus, accounting for all covariates reduced the total racial/ethnic difference for Black (−0.51 to −0.44) and Hispanic (−0.43 to −0.38) participants but differences remained significant.

### Processing Speed (Fig 1c; Table S3)

The total racial/ethnic difference in processing speed, as measured by TMT-A is 0.39 (*p* < .05) for Black and 0.47 (*p* < .05) for Hispanic participants. As expected, greater disease severity was associated with worse processing speed and attenuation of the total difference for Hispanic participants to 0.34 (*p* < .05) but not for Black participants (0.39; *p* < .05). For individual-level covariates, age and education, but not sex, were related to processing speed and attenuated the racial/ethnic difference for both Black (0.33; *p* < .05) and Hispanic (0.30; *p* < .05) participants. The presence of vascular comorbidities slightly exacerbated the racial/ethnic difference in processing speed for Black (0.36; *p* < .05) and Hispanic (0.33; *p* < .05) participants but were not related to processing speed. Adding the missing data indicator attenuated the racial/ethnic differences in processing speed for Black (0.27; *p* > .05) and Hispanic (0.30; *p* < .05) participants. Those with missing data had worse processing speed. Accounting for all covariates reduced the difference for Black, but not Hispanic, participants to non-significance.

### Cognitive Flexibility (Fig 1d; Table S4)

A significant total racial/ethnic difference in cognitive flexibility, as measured by TMT-B, was identified only for Black participants (0.60; *p* < .05). As expected, greater disease severity was associated with worse cognitive flexibility and attenuated the racial difference from 0.60 to 0.57 (*p* < .05). Age, sex and education were all associated with worse cognitive flexibility and further attenuated the difference for Black participants to 0.47 (*p* < .05). Addition of vascular comorbidities slightly exacerbated the difference to 0.48 for Black (*p* < .05) participants. Presence of diabetes was related to worse cognitive flexibility. The missing data indicator was not related to TMT-B score and did not impact the racial difference in cognitive flexibility scores for Black participants. Thus, the racial difference in cognitive flexibility remained significant for Black participants, even after considering covariates and vascular comorbidities. Although no significant difference was identified for Hispanic participants in cognitive flexibility, the total difference (0.12) was attenuated with the addition of covariates and vascular comorbidities to - 0.02 (total magnitude of .14 scaled units).

### Working Memory (Fig 1e; Table S5)

The total racial/ethnic difference in DSB, a measure of working memory, was identified only for Black participants (−0.40; *p* < .05). As expected, greater disease severity was associated with worse working memory and attenuated the racial difference to −0.35 (*p* < .05). Adding sex and education but not age further attenuated the difference for Black participants to −0.32 (*p* < .05). Sex and education, but not age, were also associated with worse working memory. The presence of vascular comorbidities was not related to working memory and their addition slightly increased the difference to −0.33 (*p* < .05) for Black participants. Adding the complete data indicator further reduced the racial difference for Black participants to non-significance (−0.26; *p* > .05). Those with missing data performed worse on working memory. Although no significant difference was identified for Hispanic participants on working memory, the total difference (− 0.09) was attenuated with the addition of covariates and vascular comorbidities to 0.09 (magnitude of 0.18 scaled units).

### Immediate Memory (Fig 1f; Table S6)

The difference in immediate verbal recall, a measure of memory, was identified only for Hispanic (−0.38; *p* < .05) participants. As expected, greater disease severity was associated with lower immediate verbal recall and attenuated the racial/ethnic difference to non-significance (−0.27; *p* > .05). Adding the covariates of age, sex and education did not attenuate the difference in a notable way but age and education were related to immediate verbal recall. The presence of vascular comorbidities slightly increased the difference in immediate verbal recall to −0.25 (*p* > .05) for Hispanic participants. The presence of diabetes was related to better immediate verbal recall. Adding the missing data indicator further reduced the racial/ethnic difference for Hispanic participants (−0.24). Those with incomplete data performed worse on immediate recall. Although no significant difference was identified for Black participants on immediate verbal recall, the total difference (−0.13) was attenuated with the addition of covariates and vascular comorbidities to −0.06.

### Delayed Memory (Fig 1g; Table S7)

We also observed a difference in delayed verbal recall for Hispanic (−0.31; *p* < .05) but not Black (−0.02; *p* > .05) participants. We identified a nearly identical pattern of racial/ethnic difference attenuation and association of individual-level factors with delayed verbal recall as for immediate verbal recall, described above. Addition of disease severity attenuated the difference to non-significance (−0.19; *p* > .05) for Hispanic and was associated with worse delayed recall. Adding age and sex had a minimal effect but addition of education further attenuated differences to −0.12 for Hispanic participants. Worse delayed recall was associated with older age and less education. Again, presence of vascular comorbidities slightly increased the difference in delayed verbal recall for Hispanic (−0.12 to −0.15) participants but remained non-significant. Presence of diabetes was related to better delayed verbal recall. Lastly, although the missing data indicator was related to worse performance on delayed recall, it did not meaningfully reduce racial/ethnic differences in delayed verbal recall for Hispanic participants. Although no significant difference was identified for Black participants on delayed verbal recall, the total difference (−0.02) was attenuated with the addition of covariates and vascular comorbidities to 0.06 (magnitude of 0.08 scaled units).

### Digit Span Forward (Fig 1h; Table S8) & Category Fluency (Fig 1i-j; Tables S9-10)

We did not observe a significant total racial/ethnic difference for attention (DSF) and category fluency (Animals and Vegetables) for Black and Hispanic participants. Although a significant total difference was not identified for these tests, the addition of disease severity, education, and the missing data indicator further reduced the estimated total difference. Attention was worse with increasing disease severity, having hypertension and missing at least one test. Category fluency measured by Vegetables was worse with increasing disease severity, older age, being male, not having diabetes, and missing at least one test. Category fluency measured by Animals was similar. Worse performance was associated with increasing disease severity, older age, being female, less education, not having diabetes, and missing at least one test.

### Differences in likelihood of impairment status in cognitive test performance

Minoritized individuals had a greater likelihood of performing below an established threshold for clinical impairment relative to their White counterparts (Fig. 2) on a few measures even when controlling for disease severity: processing speed (Black: β = 0.93, SE = 0.37, p = .013, OR = 2.54), cognitive flexibility (Black: β = 1.16, SE = 0.47, p = .014, OR = 3.20), and lexical retrieval (Black: β = 0.84, SE = 0.39, p = .030, OR = 2.32; Hispanic: β = 1.07, SE = 0.38, p = .004, OR = 2.92). In contrast to differences in neuropsychological test scores controlling for disease severity for Black participants, we did not find differences in likelihood of impairment on global cognition and working memory.

**Figure 2.**
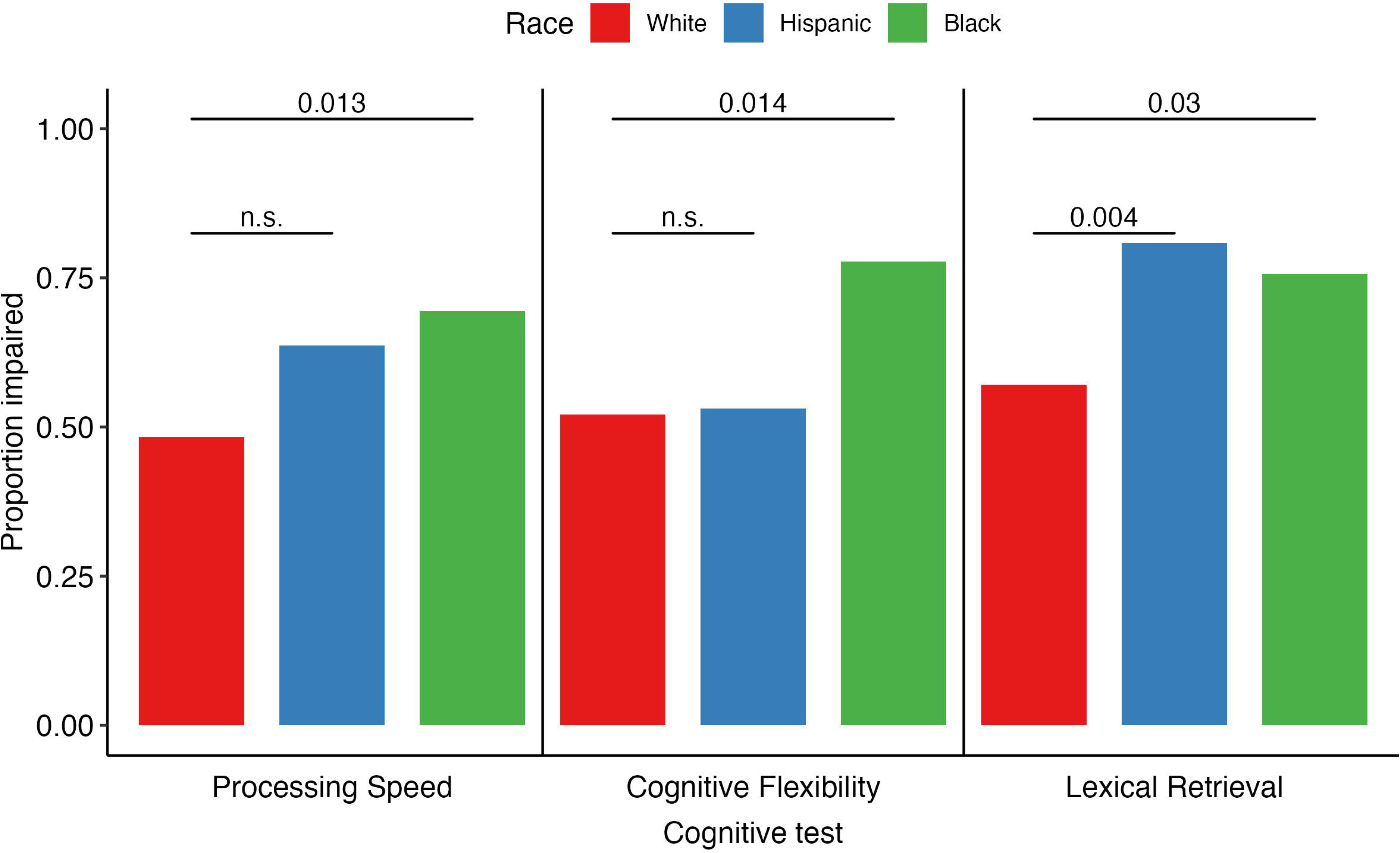
On average minoritized individuals have a higher likelihood of impairment on cognitive measures based on NACC norms adjusting for age, sex, and education.

Consistent with our lack of observing differences (after controlling for disease severity) in immediate and delayed verbal recall, attention, and category fluency, we did not identify group differences in likelihood of impairment on these tests. We did not identify any neuropsychological tests where White individuals performed significantly worse or had greater likelihood of being classified as impaired than minoritized individuals.

### Differences in reasons provided for missing cognitive test data

For all tests except global cognition, which was nearly complete for all participants included in this study, we identified significant group differences in the presence vs. absence of the test (see Table 1). As illustrated in Figure S2, Black individuals appeared more likely to be missing data due to a “Cognitive/Behavior Problem” than both White participants and, to a lesser extent, Hispanic participants.

## Discussion

We investigated whether racial and/or ethnic differences exist in neuropsychological test performance among individuals diagnosed with FTD in a large national cohort. Largely consistent with the AD^12^ and all-cause dementia^8^ literatures, we identified racial/ethnic differences in global cognition, processing speed, and lexical retrieval for Black and Hispanic individuals, working memory and cognitive flexibility for Black individuals, and immediate and delayed verbal recall for Hispanic participants. Critically, differences in test performance translated into differences in the likelihood that individuals are classified as impaired for processing speed, cognitive flexibility, lexical retrieval and delayed verbal recall. Consistent with past work^8^, we did not observe total differences for attention and category fluency for either Black or Hispanic individuals.

To our knowledge, this is the first study that examines racial/ethnic differences in neuropsychological test performance among patients with FTD. A novel and important aspect of our study is that we quantify the contribution of disease severity, common covariates (i.e., age, sex, education), and vascular comorbidities (i.e., stroke, diabetes and hypertension) to differences in cognitive performance. Critically, we implemented extensive and stringent matching between groups to account for potential systematic differences in groups on factors related to cognition. Despite these efforts, we identified significant total racial/ethnic differences for seven of the ten tests examined. Explicitly controlling for potential sources of differences in cognitive testing substantially attenuated differences for all tests and resulted in persistent differences on three tests. These findings suggest future directions for research and actions that would advance progress toward eliminating racial differences in the tools used in clinical and research settings to diagnosis, stage, and track FTD.

For most neuropsychological tests with a total difference between Hispanic and White individuals, adjusting for disease severity attenuates the difference to the point that scores, and likelihood of impairment, no longer exist. However, differences persisted for lexical retrieval and processing speed. Acculturation may account for these differences^24^. After Flores and colleagues applied normative standards developed separately for English and Spanish speakers, differences were no longer observed on a processing speed test between English and Spanish speaking Hispanic individuals^24^. Similarly, there may be cultural bias^13^, for the BNT and MINT, which reflect lexical retrieval, particularly impacting individuals with English as a second language. In the present sample, 29% of Hispanic individuals reported Spanish as their primary language. Moreover, in supplemental analyses controlling for participants’ primary language, Hispanic individuals do not perform worse than White individuals.

For Black individuals, we observed persistent differences on measures of lexical retrieval, processing speed and cognitive flexibility. Although differences are not present for Black participants on most tests of neuropsychological performance, for tests where a total difference exists (except Global Cognition), the factors considered here were inadequate in attenuating the difference to non-significance. Additionally, the presence of missing data contributed to racial differences in test performance. Qualitatively, in Figure S2, Black participants appear more likely to be missing data due to a cognitive/behavioral problem, compared to White and Hispanic participants (e.g., for DSB, Black = 14.89%, Hispanic = 7.14%, and White = 4.25% missing due to a cognitive/behavioral problem). Patients with FTD and missing neuropsychological testing likely perform worse on tests that they did complete and have more severe disease than those with complete data. However, we matched groups on disease severity and missing data and controlled for disease severity prior to adding the missing data indicator to models. Therefore, our missing data indicator captured variance not otherwise explained by other covariates or addressed through matching. Future work should investigate and test potential factors that may contribute to our observed discrepancy in data missingness.

Differences in lexical retrieval and cognitive flexibility for Black, relative to White, participants are consistent with those of prior research conducted in an all-cause dementia NACC sample^8^. This study, which included a small proportion of FTLD cases (<6.4% of Black and 14.3% of White participants), also observed that Black participants, on average, performed worse than White participants on the MoCA, TMT-A, DSB, Animals, and the MINT, when adjusting for age, sex, and years of education^8^. However, differences in disease severity and missing data across groups were not addressed in prior work, despite evidence of group differences in missing data. The overlap of our findings with that of Lennon and colleagues of racial differences in lexical retrieval and cognitive flexibility for Black participants, suggests that these differences may not be unique to FTD but due to factors that we did not capture in our analyses.

Racial/ethnic differences in neuropsychological performance were attenuated by accounting for disease severity, age, sex, education, and data missingness. However, differences persisted for some tests examined, suggesting that there are additional factors that may explain the remaining differences. In the NACC UDS, the education variable reflects quantity of education, measured in total years of education. However, there is clear evidence of the importance of education quality, not just quantity, in contributing to differences in neuropsychological test performance^14,25^. Even when Black and White older adults were matched on years of education (i.e., quantity), Black individuals obtained lower scores than White individuals, a difference that was attenuated by controlling for reading level in prior work, a proxy of education quality^14^. Furthermore, Black individuals are more likely to receive lower quality education, as measured by state-level metrics (e.g., student-teacher ratio)^26^ and school-level metrics (e.g., education attainment of teachers)^27^. Thus, we may expect that observed differences in cognitive scores would be further reduced when accounting for access to and experience of high-quality education.

Social and structural determinants of health (SSDOH)^28–31^ are additional factors that future work should examine as contributors to observed differences. It is critical to acknowledge that race, or minoritized group affiliation, is a social construct and proxy for a number of SSDOH factors^28,29^. Indeed, SSDOH explain racialized differences in cognitive outcomes among those with dementia over and above cardiovascular risk and health behaviors^32,33^: poorer cognitive function is associated with increased exposure to racism^34^, air pollution^35^, and occupational hazards^36^; and experience of poverty^37^, incarceration^38^, and poor education quality^39^. While comprehensive SSDOH measures are currently not available in the NACC UDS, UDS version 4 will introduce SSDOH measures, and thus, enable future investigation of these important factors.

In addition to SSDOH, the tools implemented to stage and diagnosis patients, and clinician interpretation of such tools may contribute to observed differences. Many standard neurocognitive tests are not valid cross-culturally due to use of culturally-bound concepts and norms based on non-representative, predominantly White, samples^40^. While alternative neuropsychological tests may reduce differences among minoritized groups and educational levels, they are not widely implemented^40^. Adding color to naming tests like the BNT and MINT reduces the association of performance and education such that better performance is less strongly associated with higher levels of education^41^. Similarly, a modified version of the TMT, Color Trails, reflects the same cognitive domains (processing speed, cognitive flexibility) but with reduced impact of culture and literacy^42^. However, none of these measures are widely applied in FTD research.

There are several limitations of currently available data to address these questions and understand how and why these cognitive differences emerge. The primary limitation of the current study is the underrepresentation of individuals from minoritized groups. A recent perspective paper authored by members of the International Society to Advance Alzheimer’s Research and Treatment FTD Professional Interest Area and Diversity and Disparities PIA discuss these barriers to inclusion in FTD research for racially and ethnically minoritized populations at length^40^. The NACC cohort diagnosed with FTD is comprised of only 2.6% Black individuals, 3.5% Hispanic individuals and 3.7% other minoritized groups. Lack of longitudinal data for minoritized individuals, particularly prior to disease onset (CDR=0) through the development of FTD (CDR > 0), hinders the ability to examine longitudinal change in cognitive domains. This lack of data also limits the ability of norms to be based on data that accurately reflects the diversity of the population. As of the December 2023 data freeze, only 175 individuals with FTD had neuropsychological data and self-identified as a race/ethnicity other than non-Hispanic White. Within this group, only 80 had complete neuropsychological data and only 4 participants had complete data before exhibiting dementia (CDR=0). Despite this, norms developed for UDS version 3 do include race^43^.

## Conclusion

Neuropsychological data in racially and ethnically minoritized populations is severely lacking in the NACC cohort diagnosed with FTD. For nearly all (7/10) neuropsychological tests examined, we observed significant total racial/ethnic differences in performance that were substantially attenuated, with 4/7 to non-significance, after adding disease severity, age, sex, education, and data missingness to models. Importantly, differences in performance, adjusted for age, sex and education and controlling for disease severity, translated to the classification of a greater proportion of minoritized, compared to White, individuals as impaired. Future efforts must focus on increasing research participation in underrepresented populations with FTD, ideally early in disease, measure factors known to contribute to racial/ethnic differences, and employ non-biased tools when available to better capture cognitive performance in all participants.

## Data Availability

Qualified researchers may obtain access to all de-identified data in the NACC registry used for this study (https://www.naccdata.org).

## Acknowledgements

We thank the participants and their families for the participation in research. The NACC database is funded by NIA/NIH Grant U24 AG072122. NACC data are contributed by the NIA-funded ADRCs: P30 AG062429 (PI James Brewer, MD, PhD), P30 AG066468 (PI Oscar Lopez, MD), P30 AG062421 (PI Bradley Hyman, MD, PhD), P30 AG066509 (PI Thomas Grabowski, MD), P30 AG066514 (PI Mary Sano, PhD), P30 AG066530 (PI Helena Chui, MD), P30 AG066507 (PI Marilyn Albert, PhD), P30 AG066444 (PI David Holtzman, MD), P30 AG066518 (PI Lisa Silbert, MD, MCR), P30 AG066512 (PI Thomas Wisniewski, MD), P30 AG066462 (PI Scott Small, MD), P30 AG072979 (PI David Wolk, MD), P30 AG072972 (PI Charles DeCarli, MD), P30 AG072976 (PI Andrew Saykin, PsyD), P30 AG072975 (PI Julie A. Schneider, MD, MS), P30 AG072978 (PI Ann McKee, MD), P30 AG072977 (PI Robert Vassar, PhD), P30 AG066519 (PI Frank LaFerla, PhD), P30 AG062677 (PI Ronald Petersen, MD, PhD), P30 AG079280 (PI Jessica Langbaum, PhD), P30 AG062422 (PI Gil Rabinovici, MD), P30 AG066511 (PI Allan Levey, MD, PhD), P30 AG072946 (PI Linda Van Eldik, PhD), P30 AG062715 (PI Sanjay Asthana, MD, FRCP), P30 AG072973 (PI Russell Swerdlow, MD), P30 AG066506 (PI Glenn Smith, PhD, ABPP), P30 AG066508 (PI Stephen Strittmatter, MD, PhD), P30 AG066515 (PI Victor Henderson, MD, MS), P30 AG072947 (PI Suzanne Craft, PhD), P30 AG072931 (PI Henry Paulson, MD, PhD), P30 AG066546 (PI Sudha Seshadri, MD), P30 AG086401 (PI Erik Roberson, MD, PhD), P30 AG086404 (PI Gary Rosenberg, MD), P20 AG068082 (PI Angela Jefferson, PhD), P30 AG072958 (PI Heather Whitson, MD), P30 AG072959 (PI James Leverenz, MD).

## Consent statement

This study involved secondary analysis of de-identified data available from naccdata.org and did not require informed consent for the present study. All participants or their caregivers provided written informed consent before data were collected at each individual ADRC, as approved by individual institutional review boards (IRBs) at each site.

