## Supplement for "Racial/Ethnic Differences in Neuropsychological Test Performance in Frontotemporal Degeneration"

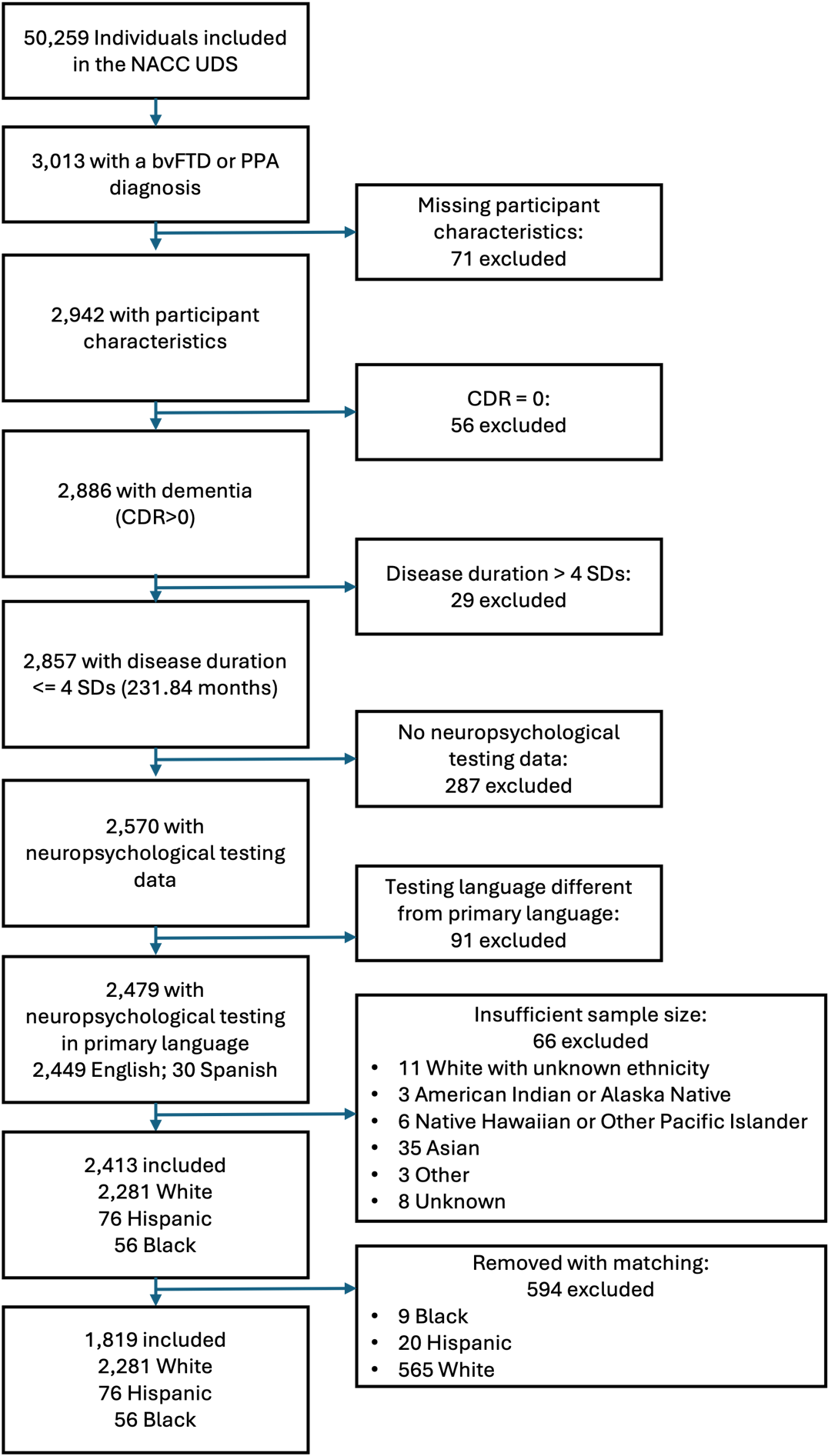

Figure S1. Flow diagram of sample selection.

**Multivariable associations for each neuropsychological test**

Table S1. Global Cognition: Multivariable associations.

| **Global Cognition^1^** | **Total difference^1^** | **Disease severity^1^** | **Age^1^** | **Sex^1^** | **Education^1^** | **Comorbidities^1^** | **Complete data^1^** |
| --- | --- | --- | --- | --- | --- | --- | --- |
| **Group** (White) |  |  |  |  |  |  |  |
| Black | -0.43* (-0.72,-0.14) | -0.27* (-0.51,-0.03) | -0.26* (-0.50,-0.02) | -0.24 (-0.48,0.01) | -0.20 (-0.44,0.04) | -0.22 (-0.46,0.01) | -0.14 (-0.37,0.08) |
| Hispanic | -0.33* (-0.60,-0.06) | -0.13 (-0.35,0.10) | -0.13 (-0.35,0.10) | -0.12 (-0.34,0.11) | -0.07 (-0.29,0.15) | -0.10 (-0.33,0.12) | -0.07 (-0.28,0.13) |
| **Disease severity** (Very mild) | |  |  |  |  |  |  |
| Mild |  | -0.26** (-0.34,-0.17) | -0.26** (-0.35,-0.18) | -0.27** (-0.36,-0.19) | -0.26** (-0.35,-0.18) | -0.27** (-0.35,-0.18) | -0.16** (-0.24,-0.08) |
| Moderate |  | -1.1** (-1.2,-0.98) | -1.1** (-1.2,-0.98) | -1.1** (-1.2,-0.99) | -1.1** (-1.2,-0.98) | -1.1** (-1.2,-0.99) | -0.74** (-0.86,-0.62) |
| Severe |  | -2.4** (-2.6,-2.2) | -2.4** (-2.6,-2.2) | -2.4** (-2.6,-2.2) | -2.4** (-2.6,-2.2) | -2.4** (-2.6,-2.2) | -1.9** (-2.1,-1.7) |
| **Age** |  |  | -0.05* (-0.09,-0.02) | -0.05* (-0.09,-0.02) | -0.06* (-0.10,-0.02) | -0.07** (-0.11,-0.03) | -0.06** (-0.10,-0.03) |
| **Sex** (Male) |  |  |  |  |  |  |  |
| Female |  |  |  | -0.15** (-0.22,-0.07) | -0.12* (-0.20,-0.04) | -0.11* (-0.19,-0.03) | -0.05 (-0.12,0.03) |
| **Education** |  |  |  |  | 0.11** (0.07,0.15) | 0.11** (0.07,0.15) | 0.10** (0.06,0.13) |
| **Stroke** |  |  |  |  |  | -0.18 (-0.54,0.17) | -0.22 (-0.55,0.10) |
| **Diabetes** |  |  |  |  |  | 0.15* (0.02,0.29) | 0.14* (0.01,0.26) |
| **Hypertension** |  |  |  |  |  | 0.09* (0.01,0.17) | 0.07 (0.00,0.15) |
| **Complete data** |  |  |  |  |  |  | -0.75** (-0.84,-0.67) |
| **(Intercept)** | 0.02 (-0.03,0.07) | 0.34** (0.28,0.39) | 0.34** (0.28,0.40) | 0.40** (0.34,0.47) | 0.38** (0.32,0.45) | 0.34** (0.26,0.41) | 0.44** (0.37,0.51) |
| R² | 0.008 | 0.311 | 0.314 | 0.319 | 0.331 | 0.336 | 0.434 |
| AIC | 5,150 | 4,492 | 4,487 | 4,475 | 4,446 | 4,439 | 4,150 |
| ^1^β (95% confidence interval) *p<0.05; **p<0.01; ***p<0.001; CI = Confidence Interval; Reference group in parentheses. | | | | | | | |

Table S2. Lexical Retrieval: Multivariable associations.

| **Lexical Retrieval^1^** | **Total difference^1^** | **Disease severity^1^** | | **Age^1^** | | **Sex^1^** | | **Education^1^** | | **Comorbidities^1^** | | **Complete data^1^** |
| --- | --- | --- | --- | --- | --- | --- | --- | --- | --- | --- | --- | --- |
| **Group** (White) |  |  | |  | |  | |  | |  | |  |
| Black | -0.51* (-0.84,-0.19) | -0.48* (-0.80,-0.16) | | -0.47* (-0.79,-0.15) | | -0.42* (-0.74,-0.11) | | -0.41* (-0.72,-0.09) | | -0.45* (-0.76,-0.13) | | -0.44* (-0.75,-0.12) |
| Hispanic | -0.43* (-0.72,-0.15) | -0.38* (-0.67,-0.10) | | -0.38* (-0.66,-0.10) | | -0.37* (-0.65,-0.09) | | -0.35* (-0.63,-0.07) | | -0.38* (-0.66,-0.10) | | -0.38* (-0.66,-0.10) |
| **Disease severity** (Very mild) | | |  | |  | |  | |  | |  | |
| Mild |  | 0.05 (-0.05,0.15) | | 0.04 (-0.06,0.14) | | 0.02 (-0.09,0.12) | | 0.02 (-0.08,0.12) | | 0.01 (-0.09,0.11) | | 0.03 (-0.08,0.13) |
| Moderate |  | -0.22* (-0.38,-0.06) | | -0.22* (-0.38,-0.06) | | -0.24* (-0.40,-0.08) | | -0.24* (-0.39,-0.08) | | -0.24* (-0.40,-0.09) | | -0.19* (-0.36,-0.03) |
| Severe |  | -1.1** (-1.4,-0.80) | | -1.1** (-1.4,-0.80) | | -1.1** (-1.4,-0.79) | | -1.1** (-1.4,-0.80) | | -1.1** (-1.4,-0.76) | | -1.0** (-1.3,-0.69) |
| **Age** |  |  | | -0.10** (-0.15,-0.06) | | -0.10** (-0.15,-0.06) | | -0.11** (-0.15,-0.06) | | -0.12** (-0.16,-0.07) | | -0.12** (-0.16,-0.07) |
| **Sex** (Male) |  |  | |  | |  | |  | |  | |  |
| Female |  |  | |  | | -0.26** (-0.35,-0.16) | | -0.25** (-0.34,-0.15) | | -0.23** (-0.33,-0.13) | | -0.22** (-0.32,-0.13) |
| **Education** |  |  | |  | |  | | 0.05 (0.00,0.09) | | 0.05 (0.00,0.09) | | 0.04 (0.00,0.09) |
| **Stroke** |  |  | |  | |  | |  | | -0.28 (-0.74,0.18) | | -0.29 (-0.75,0.17) |
| **Diabetes** |  |  | |  | |  | |  | | 0.27* (0.10,0.43) | | 0.27* (0.10,0.43) |
| **Hypertension** |  |  | |  | |  | |  | | 0.11* (0.01,0.21) | | 0.10* (0.01,0.20) |
| **Complete data** |  |  | |  | |  | |  | |  | | -0.11 (-0.23,0.00) |
| **(Intercept)** | 0.02 (-0.03,0.07) | 0.05 (-0.02,0.12) | | 0.05 (-0.01,0.12) | | 0.16** (0.09,0.24) | | 0.16** (0.08,0.24) | | 0.09* (0.00,0.18) | | 0.10* (0.02,0.19) |
| R² | 0.011 | 0.044 | | 0.054 | | 0.070 | | 0.072 | | 0.083 | | 0.085 |
| AIC | 4,728 | 4,678 | | 4,661 | | 4,635 | | 4,633 | | 4,620 | | 4,618 |
| ^1^β (95% confidence interval) *p<0.05; **p<0.01; ***p<0.001; CI = Confidence Interval; Reference group in parentheses. | | | | | | | | | | | | |

Table S3. Processing speed: Multivariable associations.

| **Processing speed^1^** | **Total difference^1^** | **Disease severity^1^** | **Age^1^** | **Sex^1^** | **Education^1^** | **Comorbidities^1^** | **Complete data^1^** |
| --- | --- | --- | --- | --- | --- | --- | --- |
| **Group** (White) |  |  |  |  |  |  |  |
| Black | 0.39* (0.06,0.72) | 0.39* (0.07,0.70) | 0.37* (0.06,0.68) | 0.36* (0.04,0.67) | 0.33* (0.02,0.65) | 0.36* (0.05,0.67) | 0.26 (-0.02,0.54) |
| Hispanic | 0.47* (0.17,0.77) | 0.34* (0.06,0.63) | 0.33* (0.05,0.62) | 0.33* (0.05,0.61) | 0.30* (0.02,0.59) | 0.33* (0.04,0.61) | 0.31* (0.05,0.56) |
| **Disease severity** (Very mild) | |  |  |  |  |  |  |
| Mild |  | 0.32** (0.22,0.42) | 0.33** (0.23,0.43) | 0.34** (0.24,0.44) | 0.33** (0.23,0.43) | 0.33** (0.23,0.43) | 0.22** (0.13,0.31) |
| Moderate |  | 0.92** (0.75,1.1) | 0.92** (0.75,1.1) | 0.92** (0.75,1.1) | 0.91** (0.74,1.1) | 0.91** (0.75,1.1) | 0.50** (0.35,0.65) |
| Severe |  | 1.3** (0.92,1.8) | 1.3** (0.87,1.7) | 1.3** (0.88,1.7) | 1.3** (0.89,1.7) | 1.3** (0.88,1.7) | 0.89** (0.51,1.3) |
| **Age** |  |  | 0.11** (0.06,0.15) | 0.11** (0.06,0.15) | 0.11** (0.06,0.16) | 0.12** (0.07,0.17) | 0.10** (0.06,0.14) |
| **Sex** (Male) |  |  |  |  |  |  |  |
| Female |  |  |  | 0.05 (-0.04,0.15) | 0.04 (-0.05,0.14) | 0.03 (-0.07,0.13) | -0.04 (-0.13,0.04) |
| **Education** |  |  |  |  | -0.05* (-0.10,-0.01) | -0.05* (-0.10,-0.01) | -0.03 (-0.07,0.01) |
| **Stroke** |  |  |  |  |  | 0.10 (-0.37,0.56) | 0.22 (-0.20,0.63) |
| **Diabetes** |  |  |  |  |  | -0.12 (-0.29,0.04) | -0.12 (-0.26,0.03) |
| **Hypertension** |  |  |  |  |  | -0.08 (-0.18,0.02) | -0.06 (-0.14,0.03) |
| **Complete data** |  |  |  |  |  |  | 1.1** (1.0,1.2) |
| **(Intercept)** | -0.02 (-0.07,0.03) | -0.25** (-0.31,-0.18) | -0.25** (-0.31,-0.18) | -0.27** (-0.35,-0.19) | -0.26** (-0.34,-0.18) | -0.22** (-0.31,-0.13) | -0.34** (-0.42,-0.26) |
| R² | 0.009 | 0.100 | 0.111 | 0.112 | 0.114 | 0.118 | 0.305 |
| AIC | 4,502 | 4,356 | 4,338 | 4,338 | 4,335 | 4,336 | 3,958 |
| ^1^β (95% confidence interval) *p<0.05; **p<0.01; ***p<0.001; CI = Confidence Interval; Reference group in parentheses. | | | | | | | |

Table S4. Cognitive flexibility: Multivariable associations.

| **Cognitive flexibility^1^** | **Total difference^1^** | **Disease severity^1^** | **Age^1^** | **Sex^1^** | **Education^1^** | **Comorbidities^1^** | **Complete data^1^** |
| --- | --- | --- | --- | --- | --- | --- | --- |
| **Group** (White) |  |  |  |  |  |  |  |
| Black | 0.60* (0.22,0.98) | 0.57* (0.20,0.94) | 0.55* (0.18,0.91) | 0.50* (0.14,0.87) | 0.47* (0.10,0.84) | 0.48* (0.11,0.85) | 0.48* (0.11,0.85) |
| Hispanic | 0.12 (-0.23,0.47) | 0.02 (-0.32,0.36) | 0.01 (-0.33,0.34) | 0.01 (-0.33,0.34) | -0.04 (-0.38,0.30) | -0.02 (-0.36,0.32) | -0.02 (-0.36,0.32) |
| **Disease severity** (Very mild) | |  |  |  |  |  |  |
| Mild |  | 0.21** (0.10,0.32) | 0.22** (0.11,0.34) | 0.24** (0.13,0.35) | 0.24** (0.12,0.35) | 0.24** (0.13,0.35) | 0.24** (0.13,0.35) |
| Moderate |  | 0.75** (0.52,0.98) | 0.72** (0.49,0.94) | 0.74** (0.51,0.96) | 0.74** (0.51,0.96) | 0.76** (0.53,0.98) | 0.76** (0.53,0.98) |
| Severe |  | 1.2** (0.62,1.8) | 1.2** (0.56,1.8) | 1.2** (0.56,1.8) | 1.2** (0.58,1.8) | 1.2** (0.56,1.8) | 1.2** (0.56,1.8) |
| **Age** |  |  | 0.18** (0.12,0.23) | 0.17** (0.12,0.23) | 0.18** (0.13,0.23) | 0.18** (0.13,0.24) | 0.18** (0.13,0.24) |
| **Sex** (Male) |  |  |  |  |  |  |  |
| Female |  |  |  | 0.14* (0.03,0.25) | 0.13* (0.02,0.24) | 0.12* (0.01,0.23) | 0.12* (0.01,0.23) |
| **Education** |  |  |  |  | -0.07* (-0.12,-0.01) | -0.07* (-0.12,-0.01) | -0.07* (-0.12,-0.01) |
| **Stroke** |  |  |  |  |  | 0.10 (-0.41,0.60) | 0.10 (-0.41,0.60) |
| **Diabetes** |  |  |  |  |  | -0.19* (-0.38,-0.01) | -0.19* (-0.38,-0.01) |
| **Hypertension** |  |  |  |  |  | -0.01 (-0.12,0.10) | -0.01 (-0.12,0.10) |
| **Complete data** |  |  |  |  |  |  | -0.04 (-0.71,0.64) |
| **(Intercept)** | -0.02 (-0.07,0.04) | -0.15** (-0.22,-0.07) | -0.15** (-0.22,-0.07) | -0.21** (-0.29,-0.12) | -0.20** (-0.28,-0.11) | -0.18** (-0.27,-0.08) | -0.18** (-0.27,-0.08) |
| R² | 0.008 | 0.052 | 0.083 | 0.088 | 0.092 | 0.096 | 0.096 |
| AIC | 3,604 | 3,551 | 3,511 | 3,507 | 3,503 | 3,504 | 3,506 |
| ^1^β (95% confidence interval) *p<0.05; **p<0.01; ***p<0.001; CI = Confidence Interval; Reference group in parentheses. | | | | | | | |

Table S5. Working memory: Multivariable associations.

| **Working memory^1^** | **Total difference^1^** | **Disease severity^1^** | **Age^1^** | **Sex^1^** | **Education^1^** | **Comorbidities^1^** | **Complete data^1^** |
| --- | --- | --- | --- | --- | --- | --- | --- |
| **Group** (White) |  |  |  |  |  |  |  |
| Black | -0.40* (-0.72,-0.08) | -0.35* (-0.66,-0.05) | -0.36* (-0.66,-0.06) | -0.34* (-0.64,-0.04) | -0.32* (-0.62,-0.01) | -0.33* (-0.63,-0.02) | -0.26 (-0.55,0.02) |
| Hispanic | -0.09 (-0.38,0.19) | 0.03 (-0.24,0.30) | 0.03 (-0.24,0.30) | 0.04 (-0.23,0.30) | 0.07 (-0.20,0.34) | 0.06 (-0.21,0.33) | 0.09 (-0.16,0.34) |
| **Disease severity** (Very mild) |  |  |  |  |  |  |  |
| Mild |  | -0.11* (-0.20,-0.01) | -0.10* (-0.20,0.00) | -0.11* (-0.21,-0.01) | -0.11* (-0.20,-0.01) | -0.11* (-0.21,-0.01) | 0.00 (-0.09,0.09) |
| Moderate |  | -0.74** (-0.90,-0.59) | -0.74** (-0.90,-0.59) | -0.75** (-0.91,-0.60) | -0.75** (-0.90,-0.60) | -0.75** (-0.90,-0.60) | -0.40** (-0.55,-0.25) |
| Severe |  | -1.6** (-1.9,-1.2) | -1.6** (-1.9,-1.2) | -1.5** (-1.9,-1.2) | -1.6** (-1.9,-1.3) | -1.6** (-1.9,-1.2) | -1.1** (-1.4,-0.80) |
| **Age** |  |  | 0.04 (0.00,0.09) | 0.04 (0.00,0.09) | 0.04 (-0.01,0.08) | 0.04 (-0.01,0.08) | 0.04* (0.00,0.09) |
| **Sex** (Male) |  |  |  |  |  |  |  |
| Female |  |  |  | -0.10* (-0.20,-0.01) | -0.09 (-0.18,0.00) | -0.09 (-0.18,0.01) | -0.03 (-0.12,0.06) |
| **Education** |  |  |  |  | 0.06* (0.02,0.11) | 0.06* (0.02,0.11) | 0.05* (0.01,0.10) |
| **Stroke** |  |  |  |  |  | 0.05 (-0.40,0.49) | -0.01 (-0.42,0.41) |
| **Diabetes** |  |  |  |  |  | 0.09 (-0.07,0.25) | 0.07 (-0.08,0.23) |
| **Hypertension** |  |  |  |  |  | 0.00 (-0.10,0.10) | -0.02 (-0.11,0.07) |
| **Complete data** |  |  |  |  |  |  | -0.81** (-0.91,-0.70) |
| **(Intercept)** | 0.01 (-0.04,0.06) | 0.16** (0.10,0.23) | 0.16** (0.09,0.23) | 0.20** (0.13,0.28) | 0.19** (0.12,0.27) | 0.19** (0.10,0.27) | 0.29** (0.21,0.37) |
| R² | 0.004 | 0.096 | 0.098 | 0.100 | 0.104 | 0.105 | 0.212 |
| AIC | 4,811 | 4,652 | 4,651 | 4,648 | 4,643 | 4,647 | 4,432 |
| ^1^β (95% confidence interval) *p<0.05; **p<0.01; ***p<0.001; CI = Confidence Interval; Reference group in parentheses. | | | | | | | |

Table S6. Immediate Memory: Multivariable associations.

| **Immediate Memory^1^** | **Total difference^1^** | **Disease severity^1^** | **Age^1^** | **Sex^1^** | **Education^1^** | **Comorbidities^1^** | **Complete data^1^** |
| --- | --- | --- | --- | --- | --- | --- | --- |
| **Group** (White) |  |  |  |  |  |  |  |
| Black | -0.13 (-0.45,0.20) | -0.09 (-0.40,0.23) | -0.08 (-0.39,0.24) | -0.07 (-0.39,0.24) | -0.04 (-0.35,0.27) | -0.08 (-0.39,0.23) | -0.05 (-0.36,0.25) |
| Hispanic | -0.38* (-0.66,-0.09) | -0.27 (-0.55,0.00) | -0.27 (-0.54,0.01) | -0.27 (-0.54,0.01) | -0.22 (-0.49,0.06) | -0.25 (-0.53,0.02) | -0.24 (-0.51,0.03) |
| **Disease severity** (Very mild) |  |  |  |  |  |  |  |
| Mild |  | -0.19** (-0.29,-0.09) | -0.19** (-0.29,-0.09) | -0.20** (-0.30,-0.10) | -0.19** (-0.29,-0.09) | -0.20** (-0.30,-0.10) | -0.13* (-0.23,-0.03) |
| Moderate |  | -0.64** (-0.80,-0.48) | -0.64** (-0.80,-0.48) | -0.64** (-0.80,-0.49) | -0.64** (-0.79,-0.48) | -0.65** (-0.81,-0.50) | -0.44** (-0.60,-0.28) |
| Severe |  | -1.2** (-1.5,-0.88) | -1.2** (-1.5,-0.88) | -1.2** (-1.5,-0.88) | -1.2** (-1.5,-0.91) | -1.2** (-1.5,-0.89) | -0.92** (-1.2,-0.60) |
| **Age** |  |  | -0.08** (-0.13,-0.03) | -0.08** (-0.13,-0.03) | -0.09** (-0.13,-0.04) | -0.10** (-0.14,-0.05) | -0.09** (-0.14,-0.04) |
| **Sex** (Male) |  |  |  |  |  |  |  |
| Female |  |  |  | -0.02 (-0.12,0.07) | -0.01 (-0.10,0.09) | 0.01 (-0.08,0.11) | 0.04 (-0.05,0.14) |
| **Education** |  |  |  |  | 0.09** (0.05,0.14) | 0.10** (0.05,0.14) | 0.09** (0.05,0.14) |
| **Stroke** |  |  |  |  |  | 0.15 (-0.30,0.61) | 0.10 (-0.35,0.55) |
| **Diabetes** |  |  |  |  |  | 0.43** (0.26,0.59) | 0.42** (0.26,0.58) |
| **Hypertension** |  |  |  |  |  | 0.03 (-0.06,0.13) | 0.02 (-0.08,0.12) |
| **Complete data** |  |  |  |  |  |  | -0.49** (-0.60,-0.38) |
| **(Intercept)** | 0.01 (-0.04,0.06) | 0.18** (0.11,0.24) | 0.18** (0.11,0.25) | 0.19** (0.11,0.27) | 0.17** (0.10,0.25) | 0.12* (0.03,0.21) | 0.18** (0.09,0.27) |
| R² | 0.004 | 0.066 | 0.072 | 0.072 | 0.081 | 0.097 | 0.135 |
| AIC | 4,756 | 4,656 | 4,646 | 4,648 | 4,634 | 4,611 | 4,540 |
| ^1^β (95% confidence interval) *p<0.05; **p<0.01; ***p<0.001; CI = Confidence Interval; Reference group in parentheses. | | | | | | | |

Table S7. Delayed Memory: Multivariable associations.

| **Delayed Memory^1^** | **Total difference^1^** | **Disease severity^1^** | **Age^1^** | **Sex^1^** | **Education^1^** | **Comorbidities^1^** | **Complete data^1^** |
| --- | --- | --- | --- | --- | --- | --- | --- |
| **Group** (White) |  |  |  |  |  |  |  |
| Black | -0.02 (-0.34,0.31) | 0.02 (-0.29,0.34) | 0.03 (-0.28,0.35) | 0.03 (-0.28,0.35) | 0.08 (-0.23,0.39) | 0.04 (-0.27,0.35) | 0.06 (-0.24,0.37) |
| Hispanic | -0.31* (-0.60,-0.02) | -0.19 (-0.47,0.09) | -0.18 (-0.46,0.09) | -0.18 (-0.46,0.09) | -0.12 (-0.39,0.16) | -0.15 (-0.43,0.13) | -0.15 (-0.42,0.13) |
| **Disease severity** (Very mild) |  |  |  |  |  |  |  |
| Mild |  | -0.33** (-0.43,-0.23) | -0.34** (-0.44,-0.24) | -0.34** (-0.44,-0.24) | -0.33** (-0.43,-0.23) | -0.34** (-0.44,-0.25) | -0.29** (-0.39,-0.19) |
| Moderate |  | -0.78** (-0.93,-0.62) | -0.78** (-0.93,-0.62) | -0.78** (-0.93,-0.62) | -0.77** (-0.92,-0.61) | -0.78** (-0.94,-0.63) | -0.63** (-0.79,-0.47) |
| Severe |  | -1.1** (-1.4,-0.81) | -1.1** (-1.4,-0.81) | -1.1** (-1.4,-0.81) | -1.2** (-1.5,-0.84) | -1.1** (-1.4,-0.82) | -0.93** (-1.2,-0.61) |
| **Age** |  |  | -0.10** (-0.15,-0.06) | -0.10** (-0.15,-0.06) | -0.11** (-0.16,-0.06) | -0.12** (-0.16,-0.07) | -0.11** (-0.16,-0.07) |
| **Sex** (Male) |  |  |  |  |  |  |  |
| Female |  |  |  | 0.00 (-0.10,0.09) | 0.02 (-0.07,0.11) | 0.04 (-0.06,0.13) | 0.06 (-0.04,0.15) |
| **Education** |  |  |  |  | 0.12** (0.08,0.17) | 0.12** (0.08,0.17) | 0.12** (0.07,0.17) |
| **Stroke** |  |  |  |  |  | 0.12 (-0.33,0.57) | 0.08 (-0.36,0.53) |
| **Diabetes** |  |  |  |  |  | 0.38** (0.22,0.54) | 0.38** (0.22,0.54) |
| **Hypertension** |  |  |  |  |  | 0.02 (-0.07,0.12) | 0.01 (-0.08,0.11) |
| **Complete data** |  |  |  |  |  |  | -0.35** (-0.46,-0.24) |
| **(Intercept)** | 0.01 (-0.04,0.06) | 0.24** (0.17,0.31) | 0.24** (0.18,0.31) | 0.24** (0.17,0.32) | 0.22** (0.15,0.30) | 0.18** (0.10,0.27) | 0.22** (0.14,0.31) |
| R² | 0.003 | 0.082 | 0.092 | 0.092 | 0.107 | 0.119 | 0.139 |
| AIC | 4,745 | 4,613 | 4,595 | 4,597 | 4,573 | 4,555 | 4,519 |
| ^1^β (95% confidence interval) *p<0.05; **p<0.01; ***p<0.001; CI = Confidence Interval; Reference group in parentheses. | | | | | | | |

Table S8. Attention: Multivariable associations.

| **Attention^1^** | **Total difference^1^** | **Disease severity^1^** | **Age^1^** | **Sex^1^** | **Education^1^** | **Comorbidities^1^** | **Complete data^1^** |
| --- | --- | --- | --- | --- | --- | --- | --- |
| **Group** (White) |  |  |  |  |  |  |  |
| Black | 0.07 (-0.25,0.39) | 0.11 (-0.20,0.42) | 0.11 (-0.20,0.42) | 0.13 (-0.18,0.44) | 0.14 (-0.17,0.45) | 0.15 (-0.16,0.47) | 0.20 (-0.11,0.50) |
| Hispanic | -0.12 (-0.40,0.16) | -0.09 (-0.37,0.18) | -0.09 (-0.37,0.18) | -0.09 (-0.36,0.19) | -0.07 (-0.34,0.21) | -0.06 (-0.34,0.21) | -0.03 (-0.30,0.23) |
| **Disease severity** (Very mild) |  |  |  |  |  |  |  |
| Mild |  | 0.10* (0.00,0.20) | 0.10* (0.00,0.20) | 0.09 (-0.01,0.20) | 0.10 (0.00,0.20) | 0.10 (-0.01,0.20) | 0.17* (0.07,0.27) |
| Moderate |  | -0.11 (-0.27,0.04) | -0.11 (-0.27,0.04) | -0.12 (-0.28,0.04) | -0.12 (-0.27,0.04) | -0.12 (-0.28,0.04) | 0.12 (-0.04,0.28) |
| Severe |  | -1.3** (-1.6,-1.0) | -1.3** (-1.6,-1.0) | -1.3** (-1.6,-0.99) | -1.3** (-1.6,-1.0) | -1.3** (-1.7,-1.0) | -1.0** (-1.3,-0.70) |
| **Age** |  |  | -0.01 (-0.06,0.04) | -0.01 (-0.06,0.04) | -0.01 (-0.06,0.03) | 0.00 (-0.05,0.04) | 0.00 (-0.04,0.05) |
| **Sex** (Male) |  |  |  |  |  |  |  |
| Female |  |  |  | -0.09 (-0.19,0.01) | -0.08 (-0.18,0.01) | -0.09 (-0.19,0.01) | -0.05 (-0.14,0.05) |
| **Education** |  |  |  |  | 0.03 (-0.01,0.08) | 0.03 (-0.02,0.08) | 0.02 (-0.02,0.07) |
| **Stroke** |  |  |  |  |  | -0.32 (-0.78,0.13) | -0.36 (-0.81,0.08) |
| **Diabetes** |  |  |  |  |  | 0.10 (-0.06,0.27) | 0.09 (-0.07,0.26) |
| **Hypertension** |  |  |  |  |  | -0.10* (-0.20,0.00) | -0.12* (-0.22,-0.02) |
| **Complete data** |  |  |  |  |  |  | -0.54** (-0.65,-0.43) |
| **(Intercept)** | 0.00 (-0.05,0.05) | 0.00 (-0.07,0.07) | 0.00 (-0.06,0.07) | 0.04 (-0.04,0.12) | 0.04 (-0.04,0.11) | 0.07 (-0.02,0.16) | 0.14* (0.05,0.23) |
| R² | 0.001 | 0.044 | 0.044 | 0.046 | 0.047 | 0.051 | 0.099 |
| AIC | 4,828 | 4,758 | 4,760 | 4,759 | 4,759 | 4,758 | 4,671 |
| ^1^β (95% confidence interval) *p<0.05; **p<0.01; ***p<0.001; CI = Confidence Interval; Reference group in parentheses. | | | | | | | |

Table S9. Category Fluency-Animals: Multivariable associations.

| **Category Fluency-Animals^1^** | **Total difference^1^** | **Disease severity^1^** | **Age^1^** | **Sex^1^** | **Education^1^** | **Comorbidities^1^** | **Complete data^1^** |
| --- | --- | --- | --- | --- | --- | --- | --- |
| **Group** (White) |  |  |  |  |  |  |  |
| Black | -0.18 (-0.50,0.13) | -0.14 (-0.44,0.16) | -0.13 (-0.43,0.17) | -0.10 (-0.40,0.20) | -0.07 (-0.37,0.23) | -0.11 (-0.41,0.19) | -0.06 (-0.34,0.23) |
| Hispanic | -0.20 (-0.48,0.07) | -0.08 (-0.34,0.19) | -0.07 (-0.33,0.19) | -0.06 (-0.32,0.20) | -0.02 (-0.29,0.24) | -0.06 (-0.32,0.20) | -0.02 (-0.27,0.23) |
| **Disease severity** (Very mild) |  |  |  |  |  |  |  |
| Mild |  | -0.17** (-0.27,-0.07) | -0.18** (-0.28,-0.08) | -0.20** (-0.30,-0.10) | -0.19** (-0.29,-0.09) | -0.20** (-0.30,-0.11) | -0.12* (-0.21,-0.03) |
| Moderate |  | -0.84** (-0.99,-0.69) | -0.84** (-0.99,-0.69) | -0.86** (-1.0,-0.71) | -0.86** (-1.0,-0.71) | -0.87** (-1.0,-0.72) | -0.59** (-0.74,-0.44) |
| Severe |  | -1.4** (-1.8,-1.1) | -1.4** (-1.8,-1.1) | -1.4** (-1.7,-1.1) | -1.4** (-1.8,-1.1) | -1.4** (-1.7,-1.1) | -1.1** (-1.4,-0.76) |
| **Age** |  |  | -0.11** (-0.16,-0.07) | -0.11** (-0.15,-0.06) | -0.11** (-0.16,-0.07) | -0.12** (-0.17,-0.08) | -0.12** (-0.16,-0.07) |
| **Sex** (Male) |  |  |  |  |  |  |  |
| Female |  |  |  | -0.21** (-0.30,-0.11) | -0.19** (-0.28,-0.10) | -0.18** (-0.27,-0.08) | -0.13* (-0.22,-0.04) |
| **Education** |  |  |  |  | 0.07* (0.03,0.12) | 0.07* (0.03,0.12) | 0.07* (0.02,0.11) |
| **Stroke** |  |  |  |  |  | -0.31 (-0.74,0.13) | -0.35 (-0.77,0.07) |
| **Diabetes** |  |  |  |  |  | 0.30** (0.14,0.46) | 0.29** (0.13,0.44) |
| **Hypertension** |  |  |  |  |  | 0.06 (-0.04,0.15) | 0.04 (-0.05,0.13) |
| **Complete data** |  |  |  |  |  |  | -0.63** (-0.74,-0.53) |
| **(Intercept)** | 0.01 (-0.04,0.06) | 0.19** (0.13,0.26) | 0.20** (0.13,0.26) | 0.28** (0.21,0.36) | 0.27** (0.20,0.35) | 0.23** (0.14,0.31) | 0.31** (0.23,0.39) |
| R² | 0.002 | 0.099 | 0.111 | 0.121 | 0.126 | 0.136 | 0.203 |
| AIC | 4,854 | 4,685 | 4,663 | 4,646 | 4,638 | 4,625 | 4,488 |
| ^1^β (95% confidence interval) *p<0.05; **p<0.01; ***p<0.001; CI = Confidence Interval; Reference group in parentheses. | | | | | | | |

Table S10. Category Fluency-Vegetables: Multivariable associations.

| **Category Fluency-Vegetables^1^** | **Total difference^1^** | **Disease severity^1^** | **Age^1^** | **Sex^1^** | **Education^1^** | **Comorbidities^1^** | **Complete data^1^** |
| --- | --- | --- | --- | --- | --- | --- | --- |
| **Group** (White) |  |  |  |  |  |  |  |
| Black | -0.03 (-0.35,0.29) | 0.01 (-0.30,0.32) | 0.02 (-0.28,0.33) | -0.01 (-0.31,0.30) | 0.00 (-0.30,0.31) | -0.03 (-0.34,0.27) | 0.01 (-0.28,0.31) |
| Hispanic | -0.25 (-0.53,0.04) | -0.13 (-0.40,0.14) | -0.13 (-0.40,0.14) | -0.14 (-0.41,0.13) | -0.13 (-0.40,0.14) | -0.17 (-0.44,0.10) | -0.15 (-0.41,0.11) |
| **Disease severity** (Very mild) |  |  |  |  |  |  |  |
| Mild |  | -0.17** (-0.27,-0.07) | -0.18** (-0.28,-0.08) | -0.17* (-0.26,-0.07) | -0.16* (-0.26,-0.06) | -0.17** (-0.27,-0.08) | -0.09 (-0.19,0.01) |
| Moderate |  | -0.75** (-0.90,-0.59) | -0.75** (-0.90,-0.59) | -0.74** (-0.89,-0.58) | -0.73** (-0.89,-0.58) | -0.74** (-0.90,-0.59) | -0.48** (-0.64,-0.32) |
| Severe |  | -1.4** (-1.7,-1.1) | -1.4** (-1.7,-1.1) | -1.4** (-1.7,-1.1) | -1.4** (-1.7,-1.1) | -1.4** (-1.7,-1.1) | -1.0** (-1.3,-0.73) |
| **Age** |  |  | -0.11** (-0.15,-0.06) | -0.11** (-0.15,-0.06) | -0.11** (-0.15,-0.06) | -0.12** (-0.17,-0.07) | -0.11** (-0.16,-0.07) |
| **Sex** (Male) |  |  |  |  |  |  |  |
| Female |  |  |  | 0.18** (0.09,0.28) | 0.19** (0.10,0.28) | 0.21** (0.11,0.30) | 0.24** (0.15,0.34) |
| **Education** |  |  |  |  | 0.03 (-0.02,0.07) | 0.03 (-0.02,0.07) | 0.02 (-0.02,0.07) |
| **Stroke** |  |  |  |  |  | -0.30 (-0.76,0.15) | -0.37 (-0.81,0.07) |
| **Diabetes** |  |  |  |  |  | 0.30** (0.14,0.46) | 0.29** (0.14,0.45) |
| **Hypertension** |  |  |  |  |  | 0.08 (-0.01,0.18) | 0.07 (-0.03,0.16) |
| **Complete data** |  |  |  |  |  |  | -0.59** (-0.70,-0.48) |
| **(Intercept)** | 0.01 (-0.04,0.06) | 0.18** (0.11,0.25) | 0.18** (0.12,0.25) | 0.11* (0.03,0.18) | 0.10* (0.02,0.18) | 0.04 (-0.04,0.13) | 0.11* (0.03,0.20) |
| R² | 0.002 | 0.084 | 0.096 | 0.103 | 0.104 | 0.115 | 0.172 |
| AIC | 4,774 | 4,635 | 4,617 | 4,604 | 4,604 | 4,589 | 4,479 |
| ^1^β (95% confidence interval) *p<0.05; **p<0.01; ***p<0.001; CI = Confidence Interval; Reference group in parentheses. | | | | | | | |

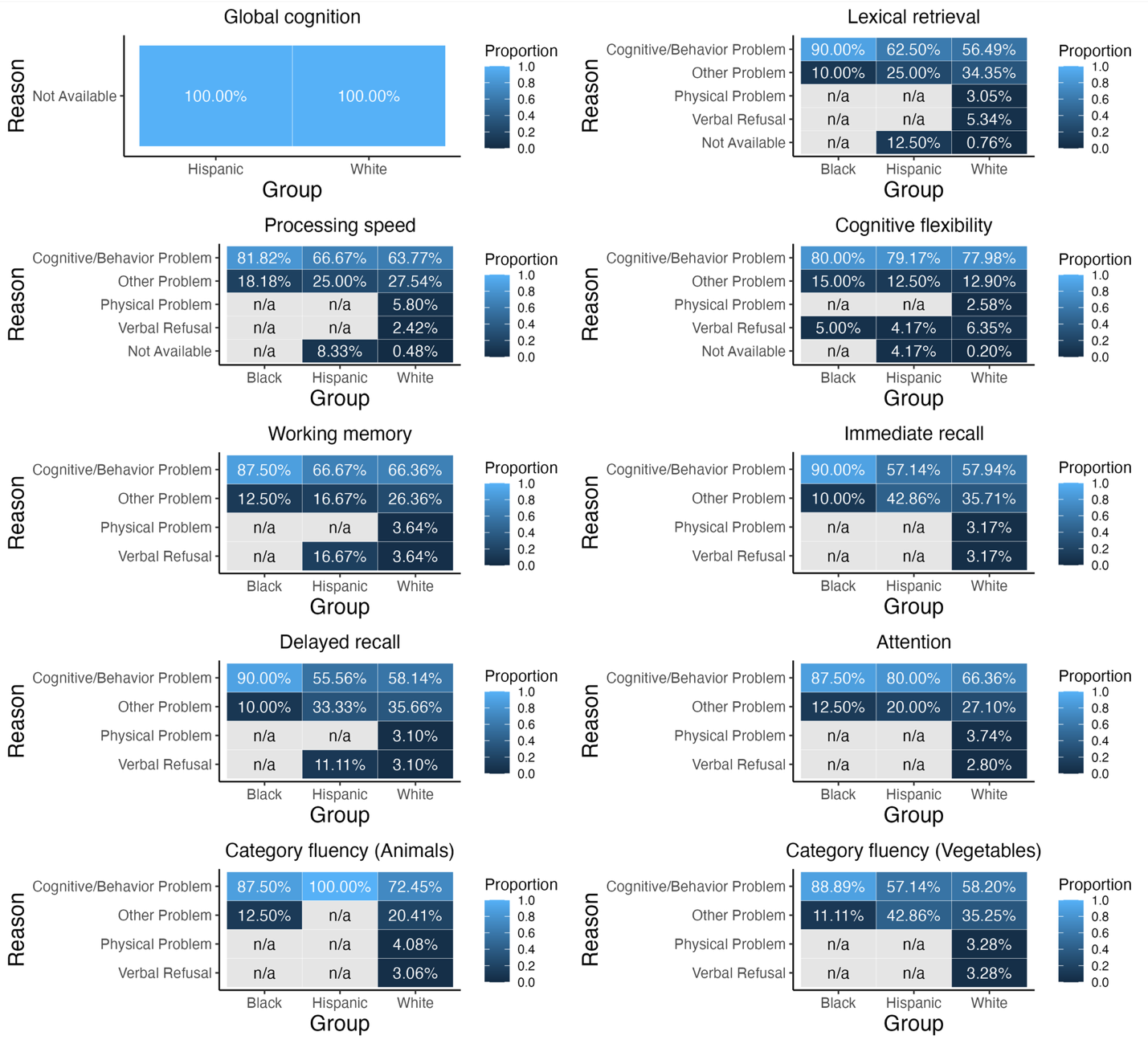

Figure S2. Data availability and reasons for missing data by group and neuropsychological test. Percentage of sample is reported in cell blocks which are color-coded by proportion.

**Within group analyses**

To further understand potential sources of racial/ethnic differences in neuropsychological test performance, we conducted analyses within racial/ethnic group, testing the effect of age, education, sex, and vascular comorbidities (i.e., presence of stroke, hypertension, and diabetes) on neuropsychological test performance accounting for disease severity.

For Black participants, worse global cognition was associated with having severe disease, being female and missing data. Worse immediate recall was associated with having moderate or severe disease. Worse delayed recall was associated with having moderate or severe disease, older age, and having complete data. Worse working memory, measured by DSB, and category fluency, measured by Animals, were associated with severe disease and being female. Worse category fluency measured by Vegetables was associated with severe disease. Worse cognitive flexibility, measured by TMT-B was associated with older age, less education, having diabetes, and having hypertension. No factors were associated with attention (DSF), processing speed (TMT-A) or lexical retrieval performance. See Table S11 for regression results.

For Hispanic participants, worse global cognition was associated with having moderate and severe disease, having diabetes, and missing data. Worse immediate recall and cognitive flexibility (TMT-B) were associated with older age. Note that all Hispanic participants with TMT-B data also had complete data. Worse attention (DSF) and category fluency measured by Animals were associated with missing data. Worse working memory (DSB) was associated with having severe disease and missing data. Worse processing speed (TMT-A) was associated with having hypertension and missing data. No factors were associated with delayed verbal recall, category fluency measured by Vegetables, or lexical retrieval performance. See Table S12 for regression results.

Consistent with analyses assessing differences in neuropsychological performance between racial/ethnic groups, for most tests, disease severity and data missingness were the most frequent predictors of test performance. This finding underscores the potential effect of minoritized individuals entering research studies, in this case the NACC UDS, with more severe disease on estimates of racial and ethnic differences. Interestingly, in these within group analyses, performance on lexical retrieval is not associated with any of the predictors, which further emphasizes our finding that the racial/ethnic differences in lexical retrieval performance remained even after accounting for these factors. Together, findings suggest that sources not accounted for here, like educational quality and cultural bias in lexical retrieval tests, may drive these effects.

Table S11. Neuropsychological test performance within Black participants

| **Predictor** | **Global cognition^1^** | **Immediate recall^1^** | **Delayed recall^1^** | **Attention^1^** | **Working memory^1^** | **Category fluency (Animals)^1^** | **Category fluency (Vegetables)^1^** | **Processing speed^1^** | **Cognitive flexibility^1^** | **Lexical retrieval^1^** |
| --- | --- | --- | --- | --- | --- | --- | --- | --- | --- | --- |
| Disease severity |  |  |  |  |  |  |  |  |  |  |
| Mild | -0.13 (-0.75,0.50) | -0.33 (-1.1,0.42) | -0.47 (-1.1,0.16) | 0.18 (-0.58,0.94) | -0.18 (-0.85,0.49) | -0.47 (-1.2,0.23) | -0.27 (-0.93,0.40) | 0.43 (-0.53,1.4) | 0.02 (-0.59,0.63) | -0.29 (-1.1,0.58) |
| Moderate | -0.62 (-1.5,0.27) | -1.7* (-3.1,-0.38) | -1.9* (-3.1,-0.76) | 1.2 (-0.17,2.5) | 0.79 (-0.40,2.0) | -0.42 (-1.7,0.82) | -1.0 (-2.2,0.10) | -0.45 (-2.2,1.3) | 0.23 (-1.7,2.2) | -0.54 (-2.1,1.0) |
| Severe | -1.8* (-2.8,-0.73) | -1.8* (-3.3,-0.32) | -1.9* (-3.2,-0.69) | -0.77 (-2.4,0.82) | -1.9* (-3.3,-0.55) | -1.7* (-3.1,-0.20) | -1.8* (-3.1,-0.43) | 0.30 (-2.2,2.8) | 0.98 (-0.49,2.5) | -1.4 (-3.1,0.39) |
| Age | -0.07 (-0.37,0.24) | -0.25 (-0.65,0.16) | -0.43* (-0.78,-0.09) | -0.09 (-0.51,0.32) | 0.02 (-0.35,0.38) | -0.14 (-0.52,0.24) | -0.21 (-0.56,0.14) | 0.35 (-0.15,0.86) | 0.49* (0.16,0.83) | -0.24 (-0.70,0.22) |
| Sex | -0.56* (-1.1,0.00) | 0.59 (-0.11,1.3) | 0.53 (-0.06,1.1) | 0.25 (-0.46,0.96) | -0.68* (-1.3,-0.06) | -0.92* (-1.6,-0.27) | -0.28 (-0.91,0.34) | 0.09 (-0.81,0.99) | 0.13 (-0.48,0.73) | -0.77 (-1.6,0.04) |
| Education | 0.16 (-0.10,0.41) | 0.05 (-0.25,0.35) | 0.11 (-0.14,0.37) | 0.06 (-0.26,0.38) | -0.03 (-0.31,0.25) | 0.12 (-0.17,0.42) | 0.04 (-0.23,0.31) | -0.16 (-0.59,0.26) | -0.49* (-0.79,-0.20) | 0.11 (-0.24,0.46) |
| Stroke | -0.25 (-1.7,1.2) | 0.81 (-1.3,2.9) | 0.01 (-1.8,1.8) | 1.5 (-0.80,3.8) | 1.8 (-0.26,3.8) | -0.78 (-2.9,1.3) | -1.4 (-3.3,0.58) | 0.19 (-2.6,3.0) | 0.65 (-1.0,2.3) | -0.97 (-3.5,1.6) |
| Diabetes | -0.13 (-0.93,0.67) | 0.23 (-0.74,1.2) | 0.27 (-0.56,1.1) | 0.86 (-0.13,1.9) | -0.13 (-1.0,0.73) | -0.29 (-1.2,0.62) | -0.55 (-1.4,0.32) | 0.40 (-0.86,1.7) | 2.1** (1.1,3.2) | -0.27 (-1.4,0.84) |
| Hypertension | 0.07 (-0.51,0.65) | -0.05 (-0.78,0.69) | -0.05 (-0.67,0.58) | -0.26 (-1.0,0.52) | -0.01 (-0.69,0.66) | 0.29 (-0.42,1.0) | 0.11 (-0.54,0.76) | -0.10 (-1.1,0.86) | -1.0* (-1.7,-0.29) | 0.09 (-0.79,0.97) |
| Data missingness | -0.71* (-1.3,-0.09) | 0.68 (-0.19,1.6) | 0.77* (0.02,1.5) | -0.81 (-1.6,0.02) | -0.41 (-1.1,0.32) | -0.70 (-1.5,0.06) | -0.39 (-1.1,0.34) | 0.42 (-0.63,1.5) | 1.5 (-0.07,3.1) | -0.14 (-1.1,0.79) |
| (Intercept) | 0.60 (-0.04,1.2) | -0.24 (-1.0,0.55) | 0.01 (-0.66,0.68) | 0.05 (-0.75,0.86) | 0.20 (-0.50,0.91) | 0.87* (0.13,1.6) | 0.66 (-0.05,1.4) | -0.08 (-1.1,0.95) | 0.38 (-0.32,1.1) | 0.30 (-0.67,1.3) |
| R² | 0.524 | 0.383 | 0.542 | 0.348 | 0.458 | 0.411 | 0.378 | 0.166 | 0.717 | 0.231 |
| ^1^β (95% confidence interval) *p<0.05; **p<0.01; ***p<0.001 | | | | | | | | | | |

Table S12. Neuropsychological test performance within Hispanic participants

| **Predictor** | **Global cognition^1^** | **Immediate recall^1^** | **Delayed recall^1^** | **Attention^1^** | **Working memory^1^** | **Category fluency (Animals)^1^** | **Category fluency (Vegetables)^1^** | **Processing speed^1^** | **Cognitive flexibility^1^** | **Lexical retrieval^1^** |
| --- | --- | --- | --- | --- | --- | --- | --- | --- | --- | --- |
| Disease severity |  |  |  |  |  |  |  |  |  |  |
| Mild | -0.07 (-0.54,0.39) | -0.06 (-0.61,0.49) | 0.03 (-0.56,0.63) | -0.27 (-0.86,0.31) | -0.47 (-0.96,0.01) | -0.02 (-0.58,0.54) | 0.06 (-0.61,0.74) | -0.21 (-0.92,0.51) | 0.00 (-0.76,0.77) | -0.13 (-0.85,0.59) |
| Moderate | -0.86* (-1.4,-0.31) | -0.18 (-0.88,0.51) | -0.45 (-1.2,0.29) | -0.10 (-0.82,0.62) | -0.58 (-1.2,0.02) | -0.67 (-1.4,0.02) | -0.33 (-1.2,0.51) | 0.11 (-0.87,1.1) | 0.16 (-0.93,1.2) | -0.29 (-1.2,0.60) |
| Severe | -1.8** (-2.6,-0.96) | -1.0 (-2.2,0.17) | -1.0 (-2.3,0.28) | -0.86 (-2.2,0.44) | -1.5* (-2.5,-0.43) | -0.97 (-2.2,0.30) | -0.90 (-2.4,0.55) | 0.23 (-1.3,1.8) | 1.8 (-0.33,3.9) | -0.65 (-2.2,0.89) |
| Age | -0.10 (-0.30,0.10) | -0.25* (-0.48,-0.01) | -0.17 (-0.43,0.08) | -0.03 (-0.29,0.22) | -0.08 (-0.28,0.12) | -0.22 (-0.47,0.02) | -0.13 (-0.41,0.16) | -0.08 (-0.39,0.24) | 0.47* (0.12,0.83) | -0.19 (-0.50,0.13) |
| Sex | 0.08 (-0.33,0.48) | 0.02 (-0.46,0.49) | 0.07 (-0.44,0.59) | 0.20 (-0.31,0.70) | 0.37 (-0.04,0.78) | 0.05 (-0.44,0.54) | 0.32 (-0.26,0.90) | -0.28 (-0.92,0.35) | -0.61 (-1.4,0.16) | 0.01 (-0.62,0.64) |
| Education | 0.07 (-0.10,0.24) | 0.03 (-0.16,0.23) | 0.07 (-0.14,0.28) | 0.04 (-0.17,0.25) | 0.06 (-0.11,0.22) | -0.01 (-0.21,0.19) | 0.00 (-0.24,0.23) | -0.05 (-0.31,0.20) | 0.07 (-0.26,0.39) | 0.16 (-0.09,0.41) |
| Diabetes | -0.84* (-1.4,-0.29) | -0.18 (-0.85,0.49) | -0.27 (-0.98,0.45) | -0.56 (-1.3,0.16) | -0.47 (-1.0,0.11) | -0.48 (-1.2,0.22) | -0.41 (-1.2,0.41) | -0.79 (-1.7,0.09) | -0.33 (-1.3,0.66) | -0.36 (-1.3,0.59) |
| Hypertension | 0.10 (-0.34,0.53) | -0.06 (-0.58,0.46) | -0.23 (-0.80,0.33) | -0.30 (-0.85,0.25) | 0.07 (-0.38,0.51) | 0.03 (-0.50,0.57) | -0.17 (-0.80,0.46) | 0.73* (0.02,1.4) | 0.42 (-0.36,1.2) | -0.24 (-0.93,0.44) |
| Data missingness | -0.92** (-1.3,-0.50) | -0.36 (-0.87,0.16) | -0.29 (-0.86,0.28) | -0.69* (-1.2,-0.15) | -1.1** (-1.6,-0.70) | -0.81* (-1.3,-0.29) | -0.58 (-1.2,0.05) | 1.8** (1.1,2.5) |  | -0.10 (-0.80,0.61) |
| (Intercept) | 0.52* (0.06,0.98) | -0.05 (-0.59,0.48) | 0.09 (-0.48,0.66) | 0.50 (-0.08,1.1) | 0.63* (0.16,1.1) | 0.34 (-0.21,0.89) | 0.05 (-0.60,0.70) | -0.12 (-0.82,0.57) | 0.05 (-0.69,0.78) | 0.02 (-0.68,0.72) |
| R² | 0.634 | 0.222 | 0.232 | 0.266 | 0.562 | 0.382 | 0.204 | 0.479 | 0.391 | 0.152 |
| ^1^β (95% confidence interval) *p<0.05; **p<0.01; ***p<0.001 | | | | | | | | | | |

**Effect of language**

In primary analyses, only individuals who were tested in their primary language were included which resulted in restriction of our analyses to examining differences across Black, Hispanic and White groups. Primary analyses were repeated including individuals who were not tested in their primary language. This allowed for the inclusion of an additional minoritized group (i.e., Asian). In these analyses, we additionally include a predictor indicating whether testing was conducted in English.

Participant Characteristics

The final sample included 37 (2.3%) Asian, 53 (2.6%) Black, 71 (3.46%) Hispanic, and 1,882 (91.7%) White individuals. Participant demographic characteristics by group are reported in Table S13. Of note, minoritized individuals were more likely to report having a primary language other than English (*p* <.001), with 54% of Hispanic and 70% of Asian participants reporting English as their primary language. In contrast, 98% of White and 94% of Black participants reported English as their primary language. Therefore, we expected that addition of primary language as a predictor of neuropsychological test performance would attenuate racial/ethnic differences for Asian and Hispanic, but not Black participants.

Table S13. Sample Characteristics

| **Characteristic** | **Overall  N = 2,053^1^** | **White  N = 1,882^1^** | **Asian  N = 47^1^** | **Black  N = 53^1^** | **Hispanic  N = 71^1^** | **p-value^2^** |
| --- | --- | --- | --- | --- | --- | --- |
| **Age (years)** | 65.26 (9.32) | 65.21 (9.27) | 65.13 (8.66) | 65.26 (10.52) | 66.63 (10.09) | 0.6 |
| **Sex (Female)** | 851 (41%) | 758 (40%) | 27 (57%) | 32 (60%) | 34 (48%) | 0.002 |
| **Education (years)** | 15.57 (2.84) | 15.65 (2.76) | 16.53 (3.17) | 14.45 (3.12) | 13.69 (3.60) | <0.001 |
| **Disease duration (years)** | 4.69 (2.92) | 4.71 (2.92) | 4.28 (3.21) | 4.45 (2.85) | 4.54 (2.90) | 0.6 |
| **Global CDR** |  |  |  |  |  | 0.14 |
| Very mild | 909 (44%) | 843 (45%) | 20 (43%) | 20 (38%) | 26 (37%) |  |
| Mild | 787 (38%) | 727 (39%) | 15 (32%) | 19 (36%) | 26 (37%) |  |
| Moderate | 269 (13%) | 236 (13%) | 10 (21%) | 9 (17%) | 14 (20%) |  |
| Severe | 88 (4.3%) | 76 (4.0%) | 2 (4.3%) | 5 (9.4%) | 5 (7.0%) |  |
| **Primary language (English)** | 1,974 (96%) | 1,853 (98%) | 33 (70%) | 50 (94%) | 38 (54%) | <0.001 |
| **NACC visit number** | 1.37 (1.00) | 1.37 (1.03) | 1.17 (0.38) | 1.28 (0.60) | 1.39 (0.92) | 0.9 |
| **Stroke** | 24 (1.2%) | 20 (1.1%) | 0 (0%) | 2 (3.8%) | 2 (2.8%) | 0.10 |
| **Diabetes** | 198 (9.6%) | 168 (8.9%) | 11 (23%) | 7 (13%) | 12 (17%) | 0.002 |
| **Hypertension** | 805 (39%) | 713 (38%) | 20 (43%) | 30 (57%) | 42 (59%) | <0.001 |
| **Clinical phenotype (bvFTD)** | 1,116 (54%) | 1,017 (54%) | 23 (49%) | 31 (58%) | 45 (63%) | 0.3 |
| **UDS version** |  |  |  |  |  |  |
| 1 | 439 (21%) | 404 (21%) | 5 (11%) | 14 (26%) | 16 (23%) |  |
| 2 | 986 (48%) | 906 (48%) | 23 (49%) | 24 (45%) | 33 (46%) |  |
| 3 | 627 (31%) | 571 (30%) | 19 (40%) | 15 (28%) | 22 (31%) |  |
| 3.2 | 1 (<0.1%) | 1 (<0.1%) | 0 (0%) | 0 (0%) | 0 (0%) |  |
| ^1^Mean (SD); n (%) | | | | | | |
| ^2^Kruskal-Wallis rank sum test; Pearson's Chi-squared test; Fisher's exact test | | | | | | |

Differences in cognitive test performance between minoritized and White groups

Regressing neuropsychological measures on race/ethnicity gives the total difference between White and minoritized (i.e., Asian, Black and Hispanic) participants in scaled units shown in the first column of Tables S14-23 and Figure S3. Given the goal of this supplementary analysis to assess the impact of language on test performance, we first add primary language (English vs. Not English) to models and focus discussion on this effect. Primary language was a significant predictor of performance on delayed recall (*p*<.05), attention (DSF; *p* <.05), working memory (DSB; *p* <.05), category fluency measured by Animals (*p* <.05), and lexical retrieval (*p* <.01).

For all minoritized participants, we identified a total racial/ethnic difference for lexical retrieval (Table S15). Lexical retrieval scores were also significantly associated with having a primary language other than English (*p*<.01). For Asian participants, addition of primary language substantially attenuated this difference (0.13 scaled units). However, none of the other predictors had much of an effect of the difference and the difference remained significant for Asian participants. For Hispanic participants, addition of primary language substantially attenuated this difference (0.27 scaled units) to non-significance (*p>*.05). Addition of disease severity further attenuated the difference by 0.05 units for Hispanic participants. For Black participants, all predictors had a small, or no, effect on the racial difference, which remained significant, consistent with primary analyses reported in the main text.

For Black and Hispanic participants, we identified a total difference for processing speed (TMT-A; Table S16). Although primary language was not associated with processing speed, its addition to the model did attenuate the performance difference for Hispanic participants by 0.09 scaled units. This difference was attenuated to non-significance with the addition of disease severity (*p*>.05). Of note, in the main text, the difference on processing speed for Hispanic participants remained significant even after all predictors were added to the model, indicating that language is a major contributing factor. As expected, primary language had minimal impact on the difference for Black participants and, consistent with analyses reported in the main text, remained significant with the addition of disease severity and other predictors.

For Hispanic participants, we identified a total difference for immediate verbal memory (Table S19). Addition of primary language attenuated this difference (0.13 scaled units) to non-significance (*p*>.05). The difference was further attenuated (0.07 units) with the addition of disease severity to the model. The only other factor that attenuated the difference for Hispanic participants was education (by 0.05 units).

For Black participants, we identified a total difference for global cognition (Table S14), working memory (DSB; Table S18) and cognitive flexibility (TMT-B; Table S17). Again, consistent with primary analyses reported in the main text, the difference for global cognition was attenuated to non-significance after the addition of disease severity and sex to the model, the difference for working memory was attenuated to non-significance after the addition of all predictors, and the difference for cognitive flexibility remained significant after accounting for all predictors. Although the total differences for global cognition, working memory, and cognitive flexibility were not significant for Asian or Hispanic participants, addition of primary language to the model reduced the group effects.

We identified no differences for attention (DSF; Table S21), category fluency measured by Animals (Table S22) or Vegetables (Table S23), and delayed verbal recall (Table S20). Addition of primary language was a significant predictor of performance on delayed recall (*p<*.05), attention (*p<*.05), and category fluency as measured by Animals (*p<*.05), where having a primary language other than English was associated with worse scores. Moreover, although there were no significant total racial/ethnic group differences on these tests, addition of the language indicator substantially reduced the non-significant difference for Asian and Hispanic participants for delayed verbal recall (Asian: -0.15 to -0.06; Hispanic: -0.25 to -0.07), attention (Asian: -0.06 to +0.03; Hispanic: -0.23 to -0.09), category fluency (Animals: Asian: -0.24 to -0.16; Hispanic: -0.14 to +0.02; Vegetables: Asian: -0.08 to -0.02; Hispanic: -0.17 to -0.05).

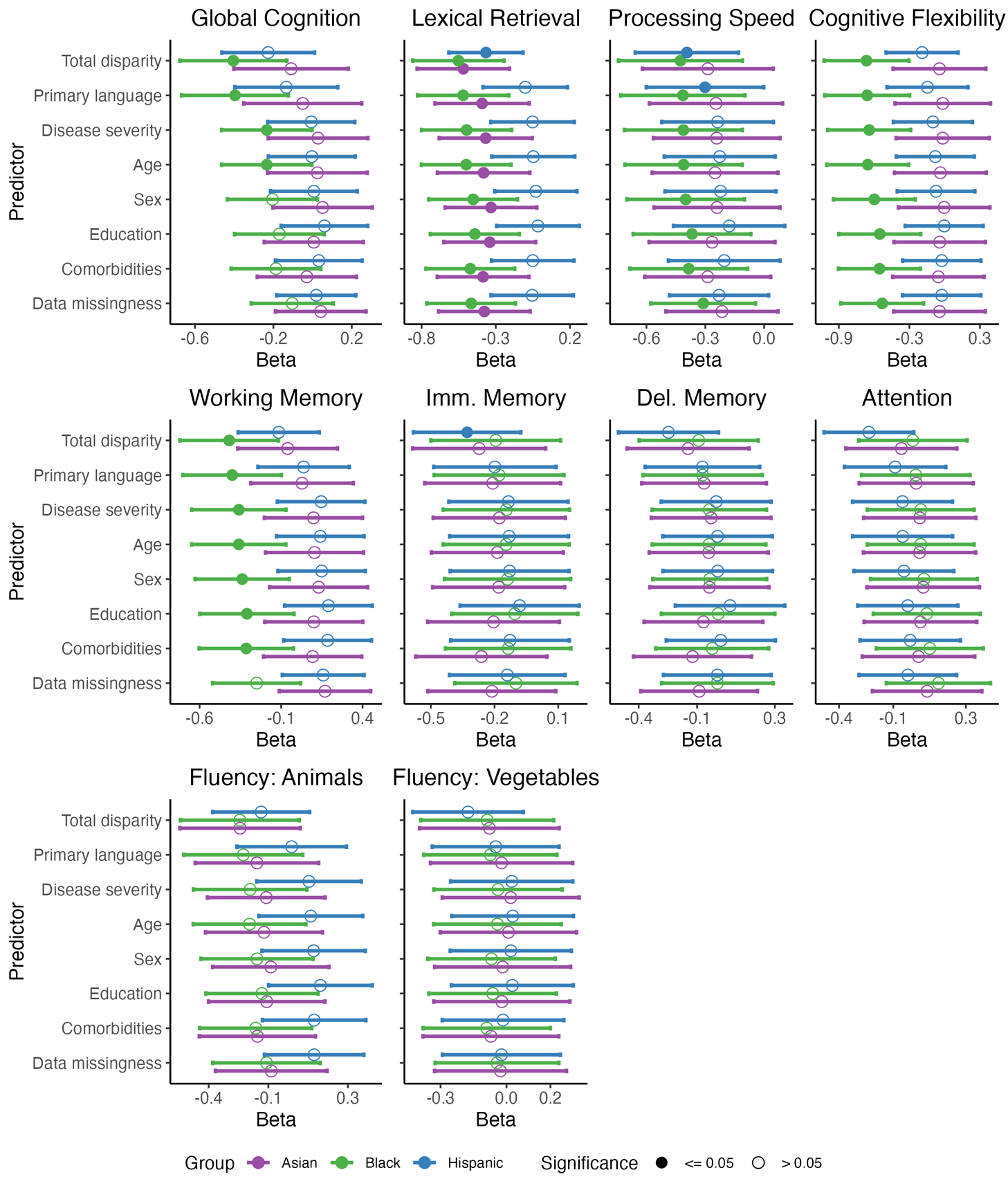
Figure S3. Estimated racial/ethnic differences by linear regression models for each neuropsychological test examined. Coefficient terms for the difference adjusting for predictors listed on the y-axis and 95% confidence intervals are displayed. Coefficients and confidence intervals were multiplied by -1 for processing speed and cognitive flexibility so that the direction of effect (higher score reflects better cognition) is the same across all tests. Abbreviations: Imm. = immediate, Del. = delayed.

Table S14. Global Cognition: Multivariable associations

| **Global Cognition^1^** | **Total difference^1^** | **Primary language^1^** | **Disease severity^1^** | **Age^1^** | **Sex^1^** | **Education^1^** | **Comorbidities^1^** | **Data missingness^1^** |
| --- | --- | --- | --- | --- | --- | --- | --- | --- |
| **Group** |  |  |  |  |  |  |  |  |
| Asian | -0.11 (-0.40,0.18) | -0.05 (-0.35,0.25) | 0.03 (-0.23,0.28) | 0.02 (-0.23,0.28) | 0.05 (-0.20,0.30) | 0.01 (-0.25,0.26) | -0.03 (-0.28,0.22) | 0.04 (-0.19,0.27) |
| Black | -0.40* (-0.68,-0.13) | -0.40* (-0.67,-0.12) | -0.23* (-0.46,0.00) | -0.23* (-0.46,0.00) | -0.20 (-0.43,0.03) | -0.17 (-0.40,0.06) | -0.19 (-0.42,0.04) | -0.10 (-0.31,0.11) |
| Hispanic | -0.23 (-0.46,0.01) | -0.13 (-0.40,0.13) | -0.01 (-0.23,0.22) | 0.00 (-0.23,0.22) | 0.01 (-0.21,0.23) | 0.06 (-0.16,0.28) | 0.03 (-0.19,0.25) | 0.02 (-0.18,0.22) |
| **Primary language** |  | -0.21 (-0.46,0.05) | -0.18 (-0.40,0.03) | -0.17 (-0.39,0.04) | -0.17 (-0.38,0.04) | -0.15 (-0.36,0.07) | -0.13 (-0.35,0.08) | -0.04 (-0.24,0.15) |
| **Disease severity** |  |  |  |  |  |  |  |  |
| Mild |  |  | -0.29** (-0.37,-0.21) | -0.30** (-0.38,-0.22) | -0.31** (-0.39,-0.23) | -0.30** (-0.38,-0.22) | -0.30** (-0.38,-0.22) | -0.16** (-0.23,-0.08) |
| Moderate |  |  | -1.0** (-1.2,-0.93) | -1.0** (-1.2,-0.93) | -1.0** (-1.2,-0.93) | -1.0** (-1.2,-0.92) | -1.0** (-1.2,-0.93) | -0.66** (-0.77,-0.55) |
| Severe |  |  | -2.3** (-2.5,-2.1) | -2.3** (-2.5,-2.1) | -2.3** (-2.5,-2.1) | -2.3** (-2.5,-2.2) | -2.3** (-2.5,-2.1) | -1.8** (-2.0,-1.6) |
| **Age** |  |  |  | -0.05* (-0.08,-0.01) | -0.05* (-0.08,-0.01) | -0.05* (-0.09,-0.02) | -0.06* (-0.10,-0.02) | -0.06* (-0.09,-0.02) |
| **Sex** |  |  |  |  | -0.15** (-0.22,-0.07) | -0.12* (-0.20,-0.05) | -0.11* (-0.19,-0.04) | -0.05 (-0.12,0.01) |
| **Education** |  |  |  |  |  | 0.10** (0.06,0.13) | 0.10** (0.06,0.13) | 0.09** (0.05,0.12) |
| **Stroke** |  |  |  |  |  |  | -0.20 (-0.54,0.13) | -0.23 (-0.53,0.08) |
| **Diabetes** |  |  |  |  |  |  | 0.15* (0.03,0.28) | 0.14* (0.03,0.26) |
| **Hypertension** |  |  |  |  |  |  | 0.07 (0.00,0.15) | 0.06 (-0.01,0.13) |
| **Data missingness** |  |  |  |  |  |  |  | -0.80** (-0.87,-0.72) |
| **(Intercept)** | 0.02 (-0.02,0.07) | 0.02 (-0.02,0.07) | 0.36** (0.31,0.42) | 0.36** (0.31,0.42) | 0.43** (0.36,0.49) | 0.41** (0.35,0.47) | 0.37** (0.30,0.44) | 0.48** (0.41,0.54) |
| R² | 0.006 | 0.007 | 0.293 | 0.295 | 0.300 | 0.309 | 0.313 | 0.428 |
| AIC | 5,812 | 5,811 | 5,122 | 5,117 | 5,104 | 5,081 | 5,074 | 4,701 |
| ^1^β (95% confidence interval) *p<0.05; **p<0.01; ***p<0.001; CI = Confidence Interval | | | | | | | | |

Table S15. Lexical Retrieval: Multivariable associations

| **Lexical Retrieval^1^** | **Total difference^1^** | **Primary language^1^** | **Disease severity^1^** | **Age^1^** | **Sex^1^** | **Education^1^** | **Comorbidities^1^** | **Data missingness^1^** |
| --- | --- | --- | --- | --- | --- | --- | --- | --- |
| **Group** |  |  |  |  |  |  |  |  |
| Asian | -0.52* (-0.83,-0.21) | -0.39* (-0.71,-0.07) | -0.37* (-0.68,-0.05) | -0.38* (-0.69,-0.07) | -0.33* (-0.64,-0.02) | -0.34* (-0.65,-0.03) | -0.39* (-0.69,-0.08) | -0.38* (-0.69,-0.07) |
| Black | -0.55** (-0.86,-0.24) | -0.52** (-0.83,-0.21) | -0.50* (-0.80,-0.19) | -0.50* (-0.80,-0.20) | -0.45* (-0.75,-0.15) | -0.44* (-0.74,-0.14) | -0.47* (-0.77,-0.17) | -0.47* (-0.76,-0.17) |
| Hispanic | -0.37* (-0.62,-0.12) | -0.10 (-0.39,0.18) | -0.05 (-0.33,0.23) | -0.05 (-0.33,0.23) | -0.03 (-0.31,0.25) | -0.02 (-0.29,0.26) | -0.05 (-0.33,0.23) | -0.05 (-0.33,0.22) |
| **Primary language** |  | -0.54** (-0.81,-0.26) | -0.54** (-0.82,-0.27) | -0.52** (-0.79,-0.25) | -0.51** (-0.78,-0.24) | -0.51** (-0.78,-0.24) | -0.49** (-0.76,-0.22) | -0.47** (-0.75,-0.20) |
| **Disease severity** |  |  |  |  |  |  |  |  |
| Mild |  |  | 0.02 (-0.08,0.11) | 0.01 (-0.09,0.10) | -0.01 (-0.11,0.08) | -0.01 (-0.11,0.08) | -0.02 (-0.11,0.08) | 0.00 (-0.10,0.10) |
| Moderate |  |  | -0.22* (-0.36,-0.07) | -0.21* (-0.35,-0.06) | -0.22* (-0.36,-0.07) | -0.21* (-0.36,-0.07) | -0.23* (-0.37,-0.08) | -0.18* (-0.33,-0.03) |
| Severe |  |  | -0.98** (-1.3,-0.68) | -0.96** (-1.3,-0.67) | -0.97** (-1.3,-0.68) | -0.98** (-1.3,-0.68) | -0.96** (-1.2,-0.67) | -0.90** (-1.2,-0.60) |
| **Age** |  |  |  | -0.11** (-0.15,-0.06) | -0.10** (-0.15,-0.06) | -0.11** (-0.15,-0.06) | -0.12** (-0.16,-0.07) | -0.12** (-0.16,-0.07) |
| **Sex** |  |  |  |  | -0.28** (-0.37,-0.19) | -0.27** (-0.36,-0.18) | -0.25** (-0.34,-0.16) | -0.25** (-0.34,-0.16) |
| **Education** |  |  |  |  |  | 0.02 (-0.02,0.07) | 0.02 (-0.02,0.07) | 0.02 (-0.02,0.07) |
| **Stroke** |  |  |  |  |  |  | -0.30 (-0.73,0.12) | -0.31 (-0.73,0.12) |
| **Diabetes** |  |  |  |  |  |  | 0.25* (0.09,0.40) | 0.24* (0.09,0.40) |
| **Hypertension** |  |  |  |  |  |  | 0.10* (0.01,0.19) | 0.10* (0.00,0.19) |
| **Data missingness** |  |  |  |  |  |  |  | -0.11* (-0.21,0.00) |
| **(Intercept)** | 0.04 (-0.01,0.08) | 0.04 (0.00,0.09) | 0.08* (0.02,0.15) | 0.08* (0.02,0.15) | 0.20** (0.13,0.28) | 0.20** (0.12,0.27) | 0.14* (0.05,0.22) | 0.15** (0.06,0.23) |
| R² | 0.016 | 0.023 | 0.049 | 0.061 | 0.079 | 0.079 | 0.089 | 0.091 |
| AIC | 5,309 | 5,297 | 5,252 | 5,231 | 5,197 | 5,197 | 5,184 | 5,182 |
| ^1^β (95% confidence interval) *p<0.05; **p<0.01; ***p<0.001; CI = Confidence Interval | | | | | | | | |

Table S16. Processing speed: Multivariable associations

| **Processing speed^1^** | **Total difference^1^** | **Primary language^1^** | **Disease severity^1^** | **Age^1^** | **Sex^1^** | **Education^1^** | **Comorbidities^1^** | **Data missingness^1^** |
| --- | --- | --- | --- | --- | --- | --- | --- | --- |
| **Group** |  |  |  |  |  |  |  |  |
| Asian | 0.29 (-0.05,0.62) | 0.25 (-0.09,0.59) | 0.24 (-0.08,0.56) | 0.25 (-0.07,0.57) | 0.24 (-0.08,0.56) | 0.27 (-0.06,0.59) | 0.29 (-0.03,0.61) | 0.22 (-0.07,0.50) |
| Black | 0.43* (0.11,0.74) | 0.41* (0.10,0.73) | 0.41* (0.11,0.71) | 0.41* (0.11,0.71) | 0.40* (0.10,0.70) | 0.37* (0.07,0.67) | 0.38* (0.08,0.68) | 0.31* (0.04,0.58) |
| Hispanic | 0.39* (0.13,0.66) | 0.30* (0.00,0.60) | 0.24 (-0.05,0.52) | 0.23 (-0.06,0.51) | 0.22 (-0.06,0.51) | 0.18 (-0.11,0.46) | 0.20 (-0.08,0.49) | 0.23 (-0.02,0.48) |
| **Primary language** |  | 0.19 (-0.11,0.49) | 0.20 (-0.09,0.48) | 0.17 (-0.11,0.45) | 0.17 (-0.11,0.45) | 0.16 (-0.12,0.45) | 0.15 (-0.13,0.44) | 0.09 (-0.16,0.34) |
| **Disease severity** |  |  |  |  |  |  |  |  |
| Mild |  |  | 0.36** (0.27,0.45) | 0.37** (0.27,0.46) | 0.37** (0.28,0.46) | 0.37** (0.27,0.46) | 0.37** (0.27,0.46) | 0.20** (0.12,0.28) |
| Moderate |  |  | 0.89** (0.74,1.0) | 0.88** (0.73,1.0) | 0.88** (0.73,1.0) | 0.87** (0.72,1.0) | 0.88** (0.73,1.0) | 0.52** (0.38,0.66) |
| Severe |  |  | 1.5** (1.2,1.9) | 1.5** (1.1,1.9) | 1.5** (1.1,1.9) | 1.5** (1.1,1.9) | 1.5** (1.1,1.9) | 1.2** (0.86,1.5) |
| **Age** |  |  |  | 0.10** (0.06,0.15) | 0.10** (0.06,0.15) | 0.11** (0.07,0.15) | 0.12** (0.07,0.16) | 0.10** (0.06,0.14) |
| **Sex** |  |  |  |  | 0.05 (-0.04,0.14) | 0.04 (-0.05,0.13) | 0.03 (-0.06,0.12) | -0.03 (-0.11,0.05) |
| **Education** |  |  |  |  |  | -0.07* (-0.11,-0.02) | -0.07* (-0.11,-0.02) | -0.05* (-0.09,-0.01) |
| **Stroke** |  |  |  |  |  |  | 0.26 (-0.17,0.68) | 0.31 (-0.07,0.69) |
| **Diabetes** |  |  |  |  |  |  | -0.08 (-0.23,0.07) | -0.09 (-0.22,0.05) |
| **Hypertension** |  |  |  |  |  |  | -0.07 (-0.17,0.02) | -0.05 (-0.14,0.03) |
| **Data missingness** |  |  |  |  |  |  |  | 1.1** (0.97,1.2) |
| **(Intercept)** | -0.03 (-0.08,0.02) | -0.03 (-0.08,0.02) | -0.28** (-0.35,-0.22) | -0.28** (-0.35,-0.22) | -0.31** (-0.38,-0.23) | -0.30** (-0.37,-0.22) | -0.26** (-0.34,-0.17) | -0.37** (-0.45,-0.30) |
| R² | 0.010 | 0.011 | 0.110 | 0.121 | 0.121 | 0.126 | 0.128 | 0.311 |
| AIC | 5,054 | 5,055 | 4,872 | 4,852 | 4,853 | 4,846 | 4,847 | 4,428 |
| ^1^β (95% confidence interval) *p<0.05; **p<0.01; ***p<0.001; CI = Confidence Interval | | | | | | | | |

Table S17. Cognitive flexibility: Multivariable associations

| **Cognitive flexibility^1^** | **Total difference^1^** | **Primary language^1^** | **Disease severity^1^** | **Age^1^** | **Sex^1^** | **Education^1^** | **Comorbidities^1^** | **Data missingness^1^** |
| --- | --- | --- | --- | --- | --- | --- | --- | --- |
| **Group** |  |  |  |  |  |  |  |  |
| Asian | 0.04 (-0.35,0.44) | 0.02 (-0.39,0.42) | 0.02 (-0.38,0.41) | 0.04 (-0.35,0.43) | 0.00 (-0.39,0.39) | 0.04 (-0.35,0.43) | 0.05 (-0.34,0.44) | 0.04 (-0.35,0.43) |
| Black | 0.66** (0.30,1.0) | 0.66** (0.30,1.0) | 0.64** (0.29,1.0) | 0.65** (0.31,1.0) | 0.60** (0.25,0.95) | 0.55* (0.20,0.90) | 0.55* (0.21,0.90) | 0.53* (0.18,0.88) |
| Hispanic | 0.19 (-0.11,0.50) | 0.15 (-0.20,0.49) | 0.10 (-0.24,0.44) | 0.08 (-0.25,0.41) | 0.07 (-0.26,0.40) | 0.00 (-0.33,0.34) | 0.02 (-0.31,0.36) | 0.02 (-0.31,0.36) |
| **Primary language** |  | 0.10 (-0.25,0.46) | 0.13 (-0.21,0.48) | 0.07 (-0.27,0.41) | 0.07 (-0.27,0.41) | 0.06 (-0.28,0.40) | 0.04 (-0.30,0.38) | 0.05 (-0.29,0.39) |
| **Disease severity** |  |  |  |  |  |  |  |  |
| Mild |  |  | 0.19** (0.08,0.30) | 0.21** (0.10,0.31) | 0.23** (0.12,0.33) | 0.22** (0.11,0.32) | 0.22** (0.11,0.33) | 0.22** (0.11,0.33) |
| Moderate |  |  | 0.72** (0.51,0.92) | 0.68** (0.48,0.89) | 0.69** (0.49,0.90) | 0.68** (0.48,0.88) | 0.70** (0.49,0.90) | 0.70** (0.49,0.90) |
| Severe |  |  | 1.3** (0.81,1.8) | 1.3** (0.79,1.7) | 1.2** (0.77,1.7) | 1.2** (0.77,1.7) | 1.2** (0.77,1.7) | 1.2** (0.76,1.7) |
| **Age** |  |  |  | 0.17** (0.12,0.22) | 0.17** (0.12,0.22) | 0.18** (0.12,0.23) | 0.18** (0.13,0.23) | 0.18** (0.13,0.23) |
| **Sex** |  |  |  |  | 0.17* (0.07,0.28) | 0.15* (0.04,0.25) | 0.14* (0.04,0.25) | 0.14* (0.03,0.25) |
| **Education** |  |  |  |  |  | -0.09** (-0.14,-0.04) | -0.09** (-0.14,-0.04) | -0.09** (-0.14,-0.04) |
| **Stroke** |  |  |  |  |  |  | 0.10 (-0.39,0.58) | 0.10 (-0.38,0.59) |
| **Diabetes** |  |  |  |  |  |  | -0.16 (-0.34,0.01) | -0.16 (-0.34,0.01) |
| **Hypertension** |  |  |  |  |  |  | 0.00 (-0.11,0.10) | 0.00 (-0.11,0.10) |
| **Data missingness** |  |  |  |  |  |  |  | 0.26 (-0.32,0.84) |
| **(Intercept)** | -0.02 (-0.08,0.03) | -0.02 (-0.08,0.03) | -0.16** (-0.23,-0.09) | -0.16** (-0.22,-0.09) | -0.23** (-0.31,-0.15) | -0.21** (-0.29,-0.13) | -0.19** (-0.29,-0.10) | -0.20** (-0.29,-0.10) |
| R² | 0.010 | 0.010 | 0.061 | 0.089 | 0.096 | 0.103 | 0.106 | 0.106 |
| AIC | 3,939 | 3,941 | 3,875 | 3,834 | 3,825 | 3,816 | 3,818 | 3,820 |
| ^1^β (95% confidence interval) *p<0.05; **p<0.01; ***p<0.001; CI = Confidence Interval | | | | | | | | |

Table S18. Working memory: Multivariable associations

| **Working memory^1^** | **Total difference^1^** | **Primary language^1^** | **Disease severity^1^** | **Age^1^** | **Sex^1^** | **Education^1^** | **Comorbidities^1^** | **Data missingness^1^** |
| --- | --- | --- | --- | --- | --- | --- | --- | --- |
| **Group** |  |  |  |  |  |  |  |  |
| Asian | -0.06 (-0.37,0.25) | 0.03 (-0.29,0.34) | 0.10 (-0.20,0.40) | 0.11 (-0.20,0.41) | 0.13 (-0.17,0.43) | 0.10 (-0.20,0.40) | 0.09 (-0.21,0.40) | 0.17 (-0.11,0.45) |
| Black | -0.42* (-0.72,-0.12) | -0.40* (-0.70,-0.10) | -0.36* (-0.65,-0.07) | -0.36* (-0.65,-0.07) | -0.34* (-0.63,-0.05) | -0.31* (-0.60,-0.02) | -0.31* (-0.60,-0.02) | -0.25 (-0.52,0.02) |
| Hispanic | -0.11 (-0.36,0.13) | 0.04 (-0.24,0.32) | 0.15 (-0.12,0.41) | 0.14 (-0.13,0.41) | 0.15 (-0.12,0.42) | 0.19 (-0.08,0.46) | 0.19 (-0.08,0.45) | 0.16 (-0.09,0.41) |
| **Primary language** |  | -0.32* (-0.60,-0.05) | -0.33* (-0.59,-0.07) | -0.34* (-0.60,-0.08) | -0.34* (-0.60,-0.08) | -0.32* (-0.58,-0.06) | -0.32* (-0.58,-0.06) | -0.23 (-0.47,0.02) |
| **Disease severity** |  |  |  |  |  |  |  |  |
| Mild |  |  | -0.14* (-0.23,-0.05) | -0.14* (-0.23,-0.04) | -0.15* (-0.24,-0.05) | -0.14* (-0.23,-0.05) | -0.14* (-0.24,-0.05) | 0.00 (-0.08,0.09) |
| Moderate |  |  | -0.71** (-0.85,-0.57) | -0.71** (-0.85,-0.57) | -0.72** (-0.86,-0.57) | -0.71** (-0.85,-0.57) | -0.71** (-0.85,-0.57) | -0.36** (-0.50,-0.23) |
| Severe |  |  | -1.5** (-1.8,-1.2) | -1.5** (-1.8,-1.2) | -1.5** (-1.8,-1.2) | -1.5** (-1.8,-1.2) | -1.5** (-1.8,-1.2) | -1.1** (-1.3,-0.80) |
| **Age** |  |  |  | 0.05* (0.00,0.09) | 0.05* (0.00,0.09) | 0.04* (0.00,0.09) | 0.04 (0.00,0.09) | 0.05* (0.01,0.09) |
| **Sex** |  |  |  |  | -0.13* (-0.21,-0.04) | -0.11* (-0.20,-0.02) | -0.11* (-0.20,-0.02) | -0.06 (-0.14,0.02) |
| **Education** |  |  |  |  |  | 0.07* (0.03,0.12) | 0.07* (0.03,0.12) | 0.07* (0.02,0.11) |
| **Stroke** |  |  |  |  |  |  | -0.02 (-0.43,0.39) | -0.04 (-0.43,0.34) |
| **Diabetes** |  |  |  |  |  |  | 0.07 (-0.08,0.22) | 0.05 (-0.09,0.19) |
| **Hypertension** |  |  |  |  |  |  | -0.01 (-0.10,0.09) | -0.02 (-0.11,0.06) |
| **Data missingness** |  |  |  |  |  |  |  | -0.83** (-0.92,-0.73) |
| **(Intercept)** | 0.01 (-0.03,0.06) | 0.02 (-0.03,0.07) | 0.18** (0.12,0.25) | 0.18** (0.12,0.25) | 0.24** (0.16,0.31) | 0.23** (0.15,0.30) | 0.22** (0.14,0.31) | 0.33** (0.25,0.41) |
| R² | 0.004 | 0.007 | 0.092 | 0.094 | 0.098 | 0.103 | 0.103 | 0.224 |
| AIC | 5,407 | 5,404 | 5,239 | 5,237 | 5,231 | 5,222 | 5,228 | 4,954 |
| ^1^β (95% confidence interval) *p<0.05; **p<0.01; ***p<0.001; CI = Confidence Interval | | | | | | | | |

Table S19. Immediate memory: Multivariable associations

| **Immediate Memory^1^** | **Total difference^1^** | **Primary language^1^** | **Disease severity^1^** | **Age^1^** | **Sex^1^** | **Education^1^** | **Comorbidities^1^** | **Data missingness^1^** |
| --- | --- | --- | --- | --- | --- | --- | --- | --- |
| **Group** |  |  |  |  |  |  |  |  |
| Asian | -0.27 (-0.59,0.04) | -0.21 (-0.53,0.11) | -0.18 (-0.49,0.13) | -0.19 (-0.50,0.12) | -0.18 (-0.49,0.13) | -0.20 (-0.52,0.11) | -0.26 (-0.57,0.05) | -0.21 (-0.51,0.09) |
| Black | -0.19 (-0.50,0.11) | -0.18 (-0.48,0.13) | -0.14 (-0.44,0.15) | -0.15 (-0.44,0.15) | -0.14 (-0.44,0.16) | -0.10 (-0.40,0.19) | -0.14 (-0.43,0.16) | -0.10 (-0.39,0.19) |
| Hispanic | -0.33* (-0.58,-0.08) | -0.20 (-0.49,0.09) | -0.13 (-0.42,0.15) | -0.13 (-0.41,0.15) | -0.13 (-0.41,0.15) | -0.08 (-0.36,0.20) | -0.13 (-0.41,0.15) | -0.14 (-0.41,0.13) |
| **Primary language** |  | -0.27 (-0.55,0.02) | -0.25 (-0.52,0.03) | -0.23 (-0.51,0.05) | -0.23 (-0.50,0.05) | -0.21 (-0.49,0.06) | -0.17 (-0.44,0.10) | -0.14 (-0.40,0.13) |
| **Disease severity** |  |  |  |  |  |  |  |  |
| Mild |  |  | -0.22** (-0.31,-0.12) | -0.22** (-0.32,-0.13) | -0.22** (-0.32,-0.13) | -0.22** (-0.31,-0.12) | -0.23** (-0.33,-0.14) | -0.13* (-0.23,-0.04) |
| Moderate |  |  | -0.58** (-0.73,-0.44) | -0.58** (-0.72,-0.43) | -0.58** (-0.72,-0.43) | -0.57** (-0.71,-0.43) | -0.59** (-0.73,-0.45) | -0.37** (-0.52,-0.22) |
| Severe |  |  | -1.1** (-1.4,-0.83) | -1.1** (-1.4,-0.82) | -1.1** (-1.4,-0.83) | -1.1** (-1.4,-0.84) | -1.1** (-1.4,-0.84) | -0.87** (-1.2,-0.58) |
| **Age** |  |  |  | -0.08** (-0.12,-0.03) | -0.08** (-0.12,-0.03) | -0.08** (-0.12,-0.04) | -0.09** (-0.13,-0.05) | -0.09** (-0.13,-0.04) |
| **Sex** |  |  |  |  | -0.04 (-0.13,0.05) | -0.03 (-0.12,0.06) | -0.01 (-0.10,0.08) | 0.02 (-0.07,0.11) |
| **Education** |  |  |  |  |  | 0.08** (0.04,0.12) | 0.08** (0.04,0.13) | 0.08** (0.04,0.12) |
| **Stroke** |  |  |  |  |  |  | 0.06 (-0.38,0.49) | 0.01 (-0.42,0.43) |
| **Diabetes** |  |  |  |  |  |  | 0.38** (0.23,0.53) | 0.37** (0.23,0.52) |
| **Hypertension** |  |  |  |  |  |  | 0.04 (-0.05,0.14) | 0.03 (-0.06,0.12) |
| **Data missingness** |  |  |  |  |  |  |  | -0.52** (-0.62,-0.42) |
| **(Intercept)** | 0.02 (-0.03,0.07) | 0.02 (-0.02,0.07) | 0.20** (0.13,0.26) | 0.20** (0.13,0.27) | 0.22** (0.14,0.29) | 0.20** (0.13,0.28) | 0.15** (0.07,0.24) | 0.21** (0.13,0.30) |
| R² | 0.006 | 0.007 | 0.062 | 0.068 | 0.068 | 0.074 | 0.088 | 0.134 |
| AIC | 5,342 | 5,341 | 5,240 | 5,231 | 5,232 | 5,221 | 5,199 | 5,104 |
| ^1^β (95% confidence interval) *p<0.05; **p<0.01; ***p<0.001; CI = Confidence Interval | | | | | | | | |

Table S20. Delayed Memory: Multivariable associations

| **Delayed Memory^1^** | **Total difference^1^** | **Primary language^1^** | **Disease severity^1^** | **Age^1^** | **Sex^1^** | **Education^1^** | **Comorbidities^1^** | **Data missingness^1^** |
| --- | --- | --- | --- | --- | --- | --- | --- | --- |
| **Group** |  |  |  |  |  |  |  |  |
| Asian | -0.15 (-0.46,0.17) | -0.06 (-0.38,0.26) | -0.03 (-0.34,0.28) | -0.04 (-0.35,0.27) | -0.04 (-0.34,0.27) | -0.07 (-0.37,0.24) | -0.12 (-0.43,0.18) | -0.09 (-0.39,0.21) |
| Black | -0.09 (-0.40,0.21) | -0.07 (-0.38,0.23) | -0.04 (-0.33,0.26) | -0.04 (-0.33,0.25) | -0.03 (-0.33,0.26) | 0.01 (-0.28,0.30) | -0.02 (-0.31,0.27) | 0.00 (-0.28,0.29) |
| Hispanic | -0.25 (-0.50,0.01) | -0.07 (-0.37,0.22) | 0.00 (-0.28,0.28) | 0.01 (-0.28,0.29) | 0.01 (-0.28,0.29) | 0.07 (-0.21,0.35) | 0.02 (-0.26,0.30) | 0.00 (-0.27,0.28) |
| **Primary language** |  | -0.35* (-0.63,-0.06) | -0.31* (-0.59,-0.04) | -0.29* (-0.57,-0.02) | -0.29* (-0.57,-0.02) | -0.27* (-0.55,0.00) | -0.23 (-0.50,0.04) | -0.20 (-0.47,0.07) |
| **Disease severity** |  |  |  |  |  |  |  |  |
| Mild |  |  | -0.36** (-0.46,-0.27) | -0.37** (-0.46,-0.28) | -0.37** (-0.47,-0.28) | -0.36** (-0.45,-0.27) | -0.38** (-0.47,-0.28) | -0.31** (-0.40,-0.21) |
| Moderate |  |  | -0.77** (-0.91,-0.62) | -0.76** (-0.90,-0.62) | -0.76** (-0.90,-0.62) | -0.75** (-0.89,-0.61) | -0.77** (-0.91,-0.63) | -0.61** (-0.76,-0.47) |
| Severe |  |  | -1.1** (-1.4,-0.79) | -1.1** (-1.4,-0.78) | -1.1** (-1.4,-0.78) | -1.1** (-1.4,-0.79) | -1.1** (-1.4,-0.80) | -0.90** (-1.2,-0.61) |
| **Age** |  |  |  | -0.10** (-0.14,-0.05) | -0.10** (-0.14,-0.05) | -0.10** (-0.15,-0.06) | -0.11** (-0.16,-0.07) | -0.11** (-0.15,-0.07) |
| **Sex** |  |  |  |  | -0.02 (-0.11,0.07) | 0.00 (-0.09,0.09) | 0.02 (-0.07,0.11) | 0.04 (-0.05,0.13) |
| **Education** |  |  |  |  |  | 0.10** (0.06,0.14) | 0.10** (0.06,0.15) | 0.10** (0.06,0.15) |
| **Stroke** |  |  |  |  |  |  | 0.04 (-0.39,0.47) | 0.01 (-0.42,0.43) |
| **Diabetes** |  |  |  |  |  |  | 0.38** (0.23,0.52) | 0.37** (0.23,0.52) |
| **Hypertension** |  |  |  |  |  |  | 0.04 (-0.05,0.13) | 0.03 (-0.06,0.12) |
| **Data missingness** |  |  |  |  |  |  |  | -0.37** (-0.47,-0.26) |
| **(Intercept)** | 0.01 (-0.03,0.06) | 0.02 (-0.03,0.06) | 0.27** (0.21,0.34) | 0.27** (0.21,0.34) | 0.28** (0.21,0.36) | 0.26** (0.19,0.34) | 0.21** (0.13,0.30) | 0.25** (0.17,0.34) |
| R² | 0.002 | 0.005 | 0.086 | 0.095 | 0.095 | 0.105 | 0.118 | 0.141 |
| AIC | 5,325 | 5,322 | 5,170 | 5,152 | 5,154 | 5,136 | 5,114 | 5,066 |
| ^1^β (95% confidence interval) *p<0.05; **p<0.01; ***p<0.001; CI = Confidence Interval | | | | | | | | |

Table S21. Attention: Multivariable associations

| **Attention^1^** | **Total difference^1^** | **Primary language^1^** | **Disease severity^1^** | **Age^1^** | **Sex^1^** | **Education^1^** | **Comorbidities^1^** | **Data missingness^1^** |
| --- | --- | --- | --- | --- | --- | --- | --- | --- |
| **Group** |  |  |  |  |  |  |  |  |
| Asian | -0.06 (-0.36,0.25) | 0.03 (-0.29,0.34) | 0.05 (-0.26,0.36) | 0.04 (-0.27,0.35) | 0.06 (-0.25,0.38) | 0.05 (-0.26,0.36) | 0.04 (-0.27,0.35) | 0.09 (-0.22,0.39) |
| Black | 0.01 (-0.29,0.31) | 0.02 (-0.28,0.32) | 0.05 (-0.24,0.35) | 0.05 (-0.24,0.35) | 0.07 (-0.23,0.36) | 0.08 (-0.21,0.38) | 0.10 (-0.19,0.40) | 0.15 (-0.14,0.44) |
| Hispanic | -0.23 (-0.48,0.01) | -0.09 (-0.37,0.19) | -0.05 (-0.33,0.23) | -0.05 (-0.32,0.23) | -0.04 (-0.32,0.23) | -0.02 (-0.30,0.26) | -0.01 (-0.28,0.27) | -0.02 (-0.29,0.25) |
| **Primary language** |  | -0.30* (-0.58,-0.03) | -0.31* (-0.58,-0.04) | -0.31* (-0.58,-0.04) | -0.31* (-0.58,-0.04) | -0.30* (-0.57,-0.03) | -0.29* (-0.56,-0.02) | -0.22 (-0.49,0.04) |
| **Disease severity** |  |  |  |  |  |  |  |  |
| Mild |  |  | 0.08 (-0.02,0.18) | 0.08 (-0.02,0.17) | 0.07 (-0.02,0.17) | 0.07 (-0.02,0.17) | 0.07 (-0.03,0.16) | 0.16** (0.07,0.26) |
| Moderate |  |  | -0.09 (-0.23,0.06) | -0.09 (-0.23,0.06) | -0.09 (-0.23,0.05) | -0.09 (-0.23,0.06) | -0.09 (-0.24,0.05) | 0.14 (-0.01,0.28) |
| Severe |  |  | -1.2** (-1.5,-0.86) | -1.2** (-1.5,-0.86) | -1.2** (-1.5,-0.87) | -1.2** (-1.5,-0.87) | -1.2** (-1.5,-0.88) | -0.91** (-1.2,-0.61) |
| **Age** |  |  |  | -0.01 (-0.05,0.04) | -0.01 (-0.05,0.04) | -0.01 (-0.05,0.04) | 0.00 (-0.04,0.05) | 0.01 (-0.04,0.05) |
| **Sex** |  |  |  |  | -0.10* (-0.19,-0.01) | -0.09* (-0.18,0.00) | -0.10* (-0.19,-0.01) | -0.06 (-0.15,0.03) |
| **Education** |  |  |  |  |  | 0.04 (-0.01,0.08) | 0.03 (-0.01,0.08) | 0.03 (-0.01,0.07) |
| **Stroke** |  |  |  |  |  |  | -0.34 (-0.77,0.08) | -0.36 (-0.77,0.05) |
| **Diabetes** |  |  |  |  |  |  | 0.15 (0.00,0.30) | 0.14 (-0.01,0.29) |
| **Hypertension** |  |  |  |  |  |  | -0.13* (-0.22,-0.03) | -0.14* (-0.23,-0.05) |
| **Data missingness** |  |  |  |  |  |  |  | -0.54** (-0.64,-0.44) |
| **(Intercept)** | 0.01 (-0.04,0.06) | 0.01 (-0.03,0.06) | 0.02 (-0.05,0.08) | 0.02 (-0.05,0.08) | 0.06 (-0.02,0.14) | 0.05 (-0.02,0.13) | 0.10* (0.01,0.18) | 0.16** (0.08,0.25) |
| R² | 0.002 | 0.004 | 0.039 | 0.039 | 0.041 | 0.043 | 0.048 | 0.100 |
| AIC | 5,423 | 5,420 | 5,359 | 5,361 | 5,358 | 5,358 | 5,352 | 5,247 |
| ^1^β (95% confidence interval) *p<0.05; **p<0.01; ***p<0.001; CI = Confidence Interval | | | | | | | | |

Table S22. Category fluency (Animals): Multivariable associations

| **Category fluency (Animals)^1^** | **Total difference^1^** | **Primary language^1^** | **Disease severity^1^** | **Age^1^** | **Sex^1^** | **Education^1^** | **Comorbidities^1^** | **Data missingness^1^** |
| --- | --- | --- | --- | --- | --- | --- | --- | --- |
| **Group** |  |  |  |  |  |  |  |  |
| Asian | -0.24 (-0.54,0.06) | -0.16 (-0.47,0.15) | -0.11 (-0.41,0.19) | -0.12 (-0.42,0.17) | -0.09 (-0.38,0.21) | -0.11 (-0.40,0.18) | -0.16 (-0.45,0.14) | -0.09 (-0.37,0.20) |
| Black | -0.24 (-0.54,0.06) | -0.23 (-0.53,0.07) | -0.19 (-0.48,0.09) | -0.19 (-0.48,0.09) | -0.16 (-0.44,0.13) | -0.13 (-0.41,0.15) | -0.16 (-0.44,0.12) | -0.11 (-0.38,0.16) |
| Hispanic | -0.14 (-0.38,0.11) | 0.02 (-0.26,0.29) | 0.10 (-0.16,0.37) | 0.11 (-0.15,0.37) | 0.13 (-0.13,0.39) | 0.16 (-0.10,0.42) | 0.13 (-0.13,0.39) | 0.13 (-0.12,0.38) |
| **Primary language** |  | -0.32* (-0.59,-0.05) | -0.30* (-0.56,-0.05) | -0.28* (-0.54,-0.03) | -0.28* (-0.53,-0.02) | -0.26* (-0.52,-0.01) | -0.24 (-0.50,0.01) | -0.17 (-0.42,0.07) |
| **Disease severity** |  |  |  |  |  |  |  |  |
| Mild |  |  | -0.21** (-0.31,-0.12) | -0.22** (-0.31,-0.13) | -0.24** (-0.33,-0.15) | -0.24** (-0.33,-0.14) | -0.24** (-0.33,-0.15) | -0.14* (-0.23,-0.05) |
| Moderate |  |  | -0.81** (-0.95,-0.67) | -0.80** (-0.94,-0.66) | -0.81** (-0.95,-0.67) | -0.80** (-0.94,-0.67) | -0.82** (-0.95,-0.68) | -0.56** (-0.69,-0.42) |
| Severe |  |  | -1.4** (-1.6,-1.1) | -1.3** (-1.6,-1.1) | -1.3** (-1.6,-1.1) | -1.4** (-1.6,-1.1) | -1.3** (-1.6,-1.1) | -1.0** (-1.3,-0.75) |
| **Age** |  |  |  | -0.11** (-0.15,-0.07) | -0.11** (-0.15,-0.07) | -0.11** (-0.16,-0.07) | -0.12** (-0.17,-0.08) | -0.12** (-0.16,-0.08) |
| **Sex** |  |  |  |  | -0.21** (-0.30,-0.13) | -0.20** (-0.29,-0.11) | -0.18** (-0.27,-0.10) | -0.15** (-0.23,-0.06) |
| **Education** |  |  |  |  |  | 0.06* (0.02,0.10) | 0.06* (0.02,0.10) | 0.05* (0.01,0.10) |
| **Stroke** |  |  |  |  |  |  | -0.35 (-0.75,0.05) | -0.37 (-0.76,0.01) |
| **Diabetes** |  |  |  |  |  |  | 0.26** (0.11,0.40) | 0.25** (0.11,0.39) |
| **Hypertension** |  |  |  |  |  |  | 0.07 (-0.02,0.16) | 0.06 (-0.03,0.14) |
| **Data missingness** |  |  |  |  |  |  |  | -0.62** (-0.72,-0.53) |
| **(Intercept)** | 0.02 (-0.03,0.06) | 0.02 (-0.03,0.07) | 0.22** (0.16,0.29) | 0.23** (0.16,0.29) | 0.32** (0.24,0.39) | 0.31** (0.23,0.38) | 0.26** (0.18,0.34) | 0.34** (0.26,0.42) |
| R² | 0.003 | 0.006 | 0.098 | 0.111 | 0.121 | 0.125 | 0.134 | 0.202 |
| AIC | 5,455 | 5,451 | 5,270 | 5,245 | 5,224 | 5,219 | 5,205 | 5,048 |
| ^1^β (95% confidence interval) *p<0.05; **p<0.01; ***p<0.001; CI = Confidence Interval | | | | | | | | |

Table S23. Category fluency (Vegetables): Multivariable associations

| **Category fluency (Vegetables)^1^** | **Total difference^1^** | **Primary language^1^** | **Disease severity^1^** | **Age^1^** | **Sex^1^** | **Education^1^** | **Comorbidities^1^** | **Data missingness^1^** |
| --- | --- | --- | --- | --- | --- | --- | --- | --- |
| **Group** |  |  |  |  |  |  |  |  |
| Asian | -0.08 (-0.40,0.24) | -0.02 (-0.35,0.30) | 0.02 (-0.29,0.33) | 0.01 (-0.30,0.32) | -0.02 (-0.33,0.29) | -0.02 (-0.33,0.29) | -0.07 (-0.38,0.24) | -0.03 (-0.33,0.27) |
| Black | -0.09 (-0.39,0.21) | -0.07 (-0.38,0.23) | -0.04 (-0.33,0.25) | -0.04 (-0.33,0.25) | -0.07 (-0.36,0.22) | -0.06 (-0.35,0.23) | -0.09 (-0.38,0.20) | -0.04 (-0.32,0.24) |
| Hispanic | -0.17 (-0.43,0.08) | -0.05 (-0.34,0.24) | 0.02 (-0.25,0.30) | 0.03 (-0.25,0.30) | 0.02 (-0.26,0.30) | 0.03 (-0.25,0.30) | -0.02 (-0.29,0.26) | -0.02 (-0.29,0.25) |
| **Primary language** |  | -0.25 (-0.54,0.03) | -0.23 (-0.51,0.04) | -0.21 (-0.48,0.07) | -0.22 (-0.49,0.06) | -0.21 (-0.49,0.06) | -0.18 (-0.45,0.09) | -0.14 (-0.41,0.12) |
| **Disease severity** |  |  |  |  |  |  |  |  |
| Mild |  |  | -0.20** (-0.29,-0.10) | -0.20** (-0.30,-0.11) | -0.19** (-0.28,-0.10) | -0.19** (-0.28,-0.09) | -0.20** (-0.29,-0.10) | -0.10* (-0.19,-0.01) |
| Moderate |  |  | -0.71** (-0.85,-0.56) | -0.70** (-0.84,-0.56) | -0.69** (-0.84,-0.55) | -0.69** (-0.84,-0.55) | -0.71** (-0.85,-0.57) | -0.48** (-0.62,-0.33) |
| Severe |  |  | -1.3** (-1.6,-1.0) | -1.3** (-1.6,-0.99) | -1.3** (-1.6,-0.99) | -1.3** (-1.6,-0.99) | -1.3** (-1.6,-0.98) | -0.99** (-1.3,-0.70) |
| **Age** |  |  |  | -0.10** (-0.14,-0.05) | -0.10** (-0.14,-0.05) | -0.10** (-0.14,-0.05) | -0.11** (-0.15,-0.06) | -0.10** (-0.14,-0.06) |
| **Sex** |  |  |  |  | 0.16** (0.07,0.25) | 0.17** (0.08,0.25) | 0.18** (0.09,0.27) | 0.21** (0.13,0.30) |
| **Education** |  |  |  |  |  | 0.01 (-0.03,0.06) | 0.01 (-0.03,0.06) | 0.01 (-0.03,0.05) |
| **Stroke** |  |  |  |  |  |  | -0.35 (-0.78,0.08) | -0.40 (-0.82,0.02) |
| **Diabetes** |  |  |  |  |  |  | 0.28** (0.13,0.43) | 0.28** (0.13,0.42) |
| **Hypertension** |  |  |  |  |  |  | 0.08 (-0.02,0.17) | 0.06 (-0.03,0.15) |
| **Data missingness** |  |  |  |  |  |  |  | -0.54** (-0.65,-0.44) |
| **(Intercept)** | 0.01 (-0.04,0.06) | 0.01 (-0.03,0.06) | 0.20** (0.13,0.26) | 0.20** (0.13,0.26) | 0.13** (0.05,0.20) | 0.13* (0.05,0.20) | 0.07 (-0.01,0.16) | 0.14* (0.05,0.22) |
| R² | 0.001 | 0.003 | 0.078 | 0.087 | 0.093 | 0.093 | 0.104 | 0.155 |
| AIC | 5,353 | 5,352 | 5,211 | 5,194 | 5,183 | 5,185 | 5,169 | 5,060 |
| ^1^β (95% confidence interval) *p<0.05; **p<0.01; ***p<0.001; CI = Confidence Interval | | | | | | | | |
